# Synthesizing multidimensional clinical profiles from published Kaplan–Meier images

**DOI:** 10.64898/2026.03.17.26348584

**Authors:** Zheqing Zhu, Fangfang Shen, Yuhua Qian, Jieting Wang

## Abstract

Strict privacy regulations restrict access to individual-level clinical trial data, confining secondary evidence synthesis to isolated one-dimensional Kaplan-Meier summaries. Inferring unreported joint distributions from these independent marginals constitutes an ill-posed inverse problem that limits multivariable precision oncology. Conventional algorithms default to the maximum entropy (MaxEnt) prior, which forces uncalibrated conditional independence upon highly collinear variables. This dismantles inherent informational dependencies, yielding unrealistic, “off-manifold” profiles that fundamentally misspecify the data-generating process and distort multivariable credit allocation. Here, we introduce MD-JoPiGo, an open-source computational framework that overcomes this structural unidentifiability by constraining high-dimensional integration within deterministic Fréchet bounds. Drawing on partial identification principles, the algorithm employs quadratically penalized maximum entropy combined with simulated annealing to anchor point estimations within the theoretically observable Fréchet gap. Through causal topology simulations, we demonstrate that the uncalibrated MaxEnt prior suffices for weakly correlated predictors (*r* ≤ 0.4) but requires structural calibration to prevent systematic error under high collinearity (*r* ≥ 0.6). This topological constraint functions as a variance firewall, ensuring that structural uncertainty and prognostic misallocation remain strictly localized. Extensive Monte Carlo simulations and empirical benchmark analyses—including lung cancer (*n* = 228), stage III colon cancer (*N* = 929), and CheckMate 227—demonstrate that the framework successfully recovers latent intersectional treatment effects. By unifying topological inference with data generation, this method translates fragmented public evidence into realistic *in silico* cohorts, supplying the multivariable resolution required for cross-study benchmarking and privacy-preserving Synthetic Control Arm (SCA) construction.

## Main

Characterizing the heterogeneity of treatment effects (HTE) is central to precision oncology, requiring high-resolution multivariable patient profiles to identify intersecting clinical features that drive survival outcomes^1–3^. However, strict privacy regulations restrict individual-level clinical trial data to proprietary databases, as releasing high-dimensional joint profiles poses severe risks of patient re-identification^4–6^. To accommodate these privacy mandates and publication constraints, randomized controlled trials typically reduce data dimensionality, publishing patient outcomes as isolated, one-dimensional (1D) Kaplan-Meier (KM) survival summaries^7–9^. This statistical marginalization obscures the intersectional probabilities required for tailored therapeutic decisions^10^. Attempting to estimate treatment efficacy across patient subcohorts from these aggregate summaries without accounting for within-trial interactions introduces severe ecological bias^11,12^. Indeed, reconstructing pseudo-individual patient data (IPD) from marginal statistics hinges entirely on specifying the joint distribution and its underlying, often unobserved, correlation structures^13^. Consequently, inferring the underlying joint distribution from independent marginals constitutes a fundamental, ill-posed inverse problem that currently limits evidence synthesis^14^.

Lacking access to the true joint distribution, conventional synthesis Lacking access to the true joint distribution, conventional synthesis frameworks—such as population-adjusted matching and marginal reweighting^15,16^—address this structural unidentifiability by implicitly or explicitly defaulting to the maximum entropy (MaxEnt) prior. Constrained solely by isolated 1D marginals, this approach mathematically forces statistical independence between clinical features. While computationally convenient, this assumption fundamentally conflicts with the biological and clinical realities of oncology trials, where covariates often exhibit strong multicollinearity. Applying an uncalibrated MaxEnt prior upon highly correlated variables (*r* ≥ 0.6) erases their inherent informational dependencies, generating unrealistic, “off-manifold” multivariable profiles that introduce substantial structural misspecification. Ultimately, this structural distortion nonlinearly amplifies hazard errors and systematically biases the reconstructed prognostic weights—a vulnerability analogous to the feature-attribution crisis in machine learning, where marginal approximations of correlated variables fundamentally distort multivariable credit allocation^17^ and exacerbate the spurious correlations that undermine survival models under distribution shifts^18^.

To resolve this structural bottleneck, we introduce MD-JoPiGo, an open-source computational framework that overcomes unidentifiability by constraining high-dimensional integration within deterministic bounds. Rather than relying on unconstrained entropy maximization, MD-JoPiGo regularizes the synthesis process by subjecting it to strict topological boundaries. Grounded in partial identification^19^, the algorithm rejects unverifiable point assumptions. Instead, it employs quadratically penalized maximum entropy combined with simulated annealing^20^ to anchor the reconstructed joint distribution strictly within the theoretically observable Fréchet gap^21^. By mapping the exact mathematical boundaries of the joint distribution space, this approach prevents entropy-driven drift and allows for the dynamic integration of minimal causal priors to correct localized misallocation.

As the first computational framework capable of multi-dimensional survival synthesis, MD-JoPiGo establishes the fundamental operational constraints for joint survival manifolds. Through causal topology simulations, we quantify specific operational thresholds. For weakly correlated predictors (*r* ≤ 0.4), the naive MaxEnt prior suffices. Under high collinearity (*r* ≥ 0.6), structural calibration—guided by Directed Acyclic Graphs (DAGs)^22,23^—is required to prevent inferential collapse. Within this framework, the topological constraint functions as a novel structural variance firewall, ensuring that prognostic misallocation remains tightly localized and does not propagate across the broader multivariable network^24^.

We validate this deterministic synthesis across Monte Carlo benchmarks and empirical oncology cohorts—including lung cancer (*N* = 228^25^), stage III colon cancer (*N* = 929^26^), and fragmented reports from the CheckMate 227 trial^27,28^—confirming that MD-JoPiGo recovers latent intersectional treatment effects. By deterministically bounding the unidentifiable space, this framework translates fragmented public evidence into privacy-preserving, synthetic individual patient data (IPD), establishing a computational foundation for cross-study benchmarking and synthetic control arm construction.

## Results

### Performance and adaptation of MD-JoPiGo

Multivariable reconstruction from 1D-IPD is fundamentally governed by the underlying causal structure of the clinical variables. To evaluate robustness across distinct data-generating mechanisms, we simulated three canonical causal topologies using directed acyclic graphs (DAGs)^24^: (1) parallel independence (Scenario A), where predictors exert independent survival effects; (2) mediation (Scenario B), where one predictor causes another, inducing inherent correlation; and (3) collider stratification (Scenario C), where independent variables jointly determine trial enrollment, inducing spurious correlation (Berkson’s paradox). Using Monte Carlo simulations (200 iterations; cohort *N* = 200) across these DAGs (Fig. 2a–c), we compared reconstructed multivariable hazard ratios with ground-truth profiles to define the operational limits of the framework.

**Fig. 1.**
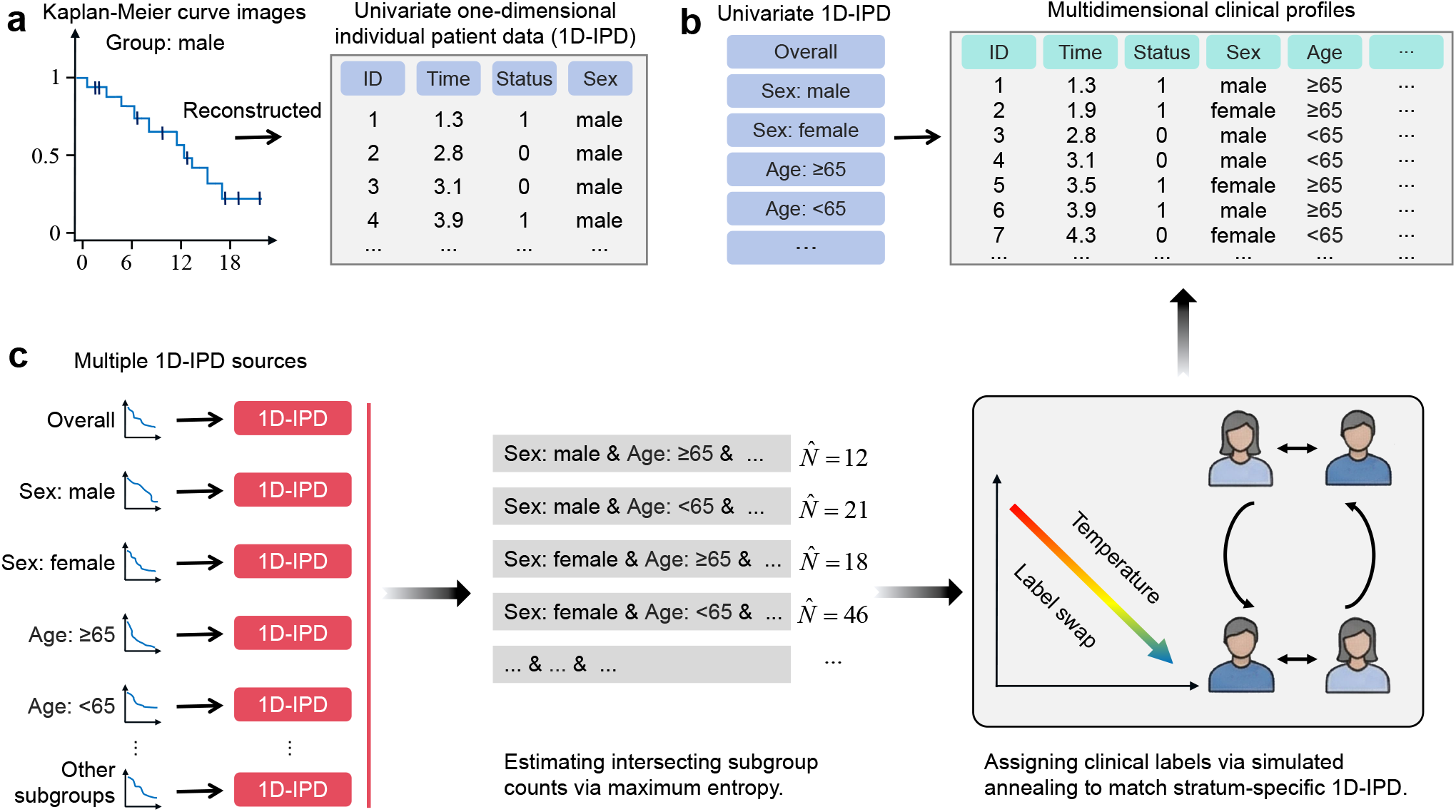
Schematic of the MD-JoPiGo framework for synthesizing multidimensional clinical profiles from published Kaplan–Meier curves. a. Reconstruction of one-dimensional individual patient data (1D-IPD). Published Kaplan–Meier (KM) curves (e.g., the male subgroup) are digitized to extract individual survival times and event statuses, yielding 1D-IPD that map survival outcomes to isolated clinical dimensions. **b. Transition from fragmented 1D-IPD to a unified multidimensional cohort**. Separate 1D-IPD sets (e.g., overall, sex-, and age-stratified populations) are integrated into a single unified cohort, assigning each patient a complete set of concurrent clinical labels. **c. The two-stage optimization pipeline**. First, maximum entropy optimization estimates the unobserved joint subgroup frequencies (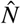; e.g., female patients aged *<* 65) from the available 1D-IPD marginals. Second, simulated annealing assigns these combinatorial labels to individual patients. A temperature-controlled label-swapping mechanism iteratively optimizes this assignment until the synthesized multivariable profiles reproduce the original stratum-specific survival trajectories.

**Fig. 2.**
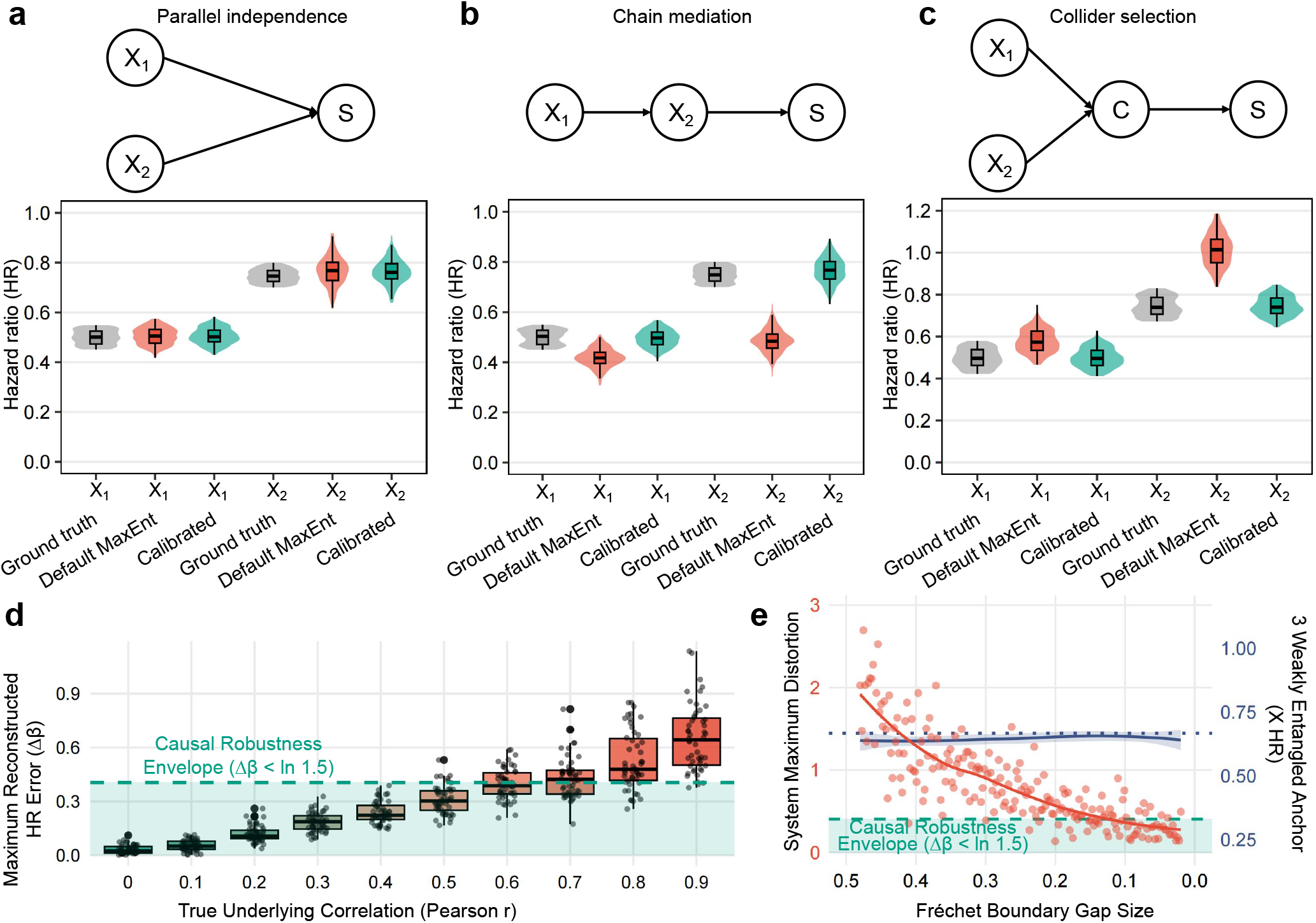
Reconstruction of multivariable hazard ratios across distinct causal topologies using MD-JoPiGo. Monte Carlo simulations (200 iterations; cohort *N* = 200) were conducted. All presented hazard ratios (HRs) are multivariable-adjusted, estimated via Cox proportional hazards models incorporating both *X*_1_ and *X*_2_. In the directed acyclic graphs (DAGs), *S* denotes survival outcome, and *C* denotes a collider representing selection criteria. **(a) Parallel independence:** Under covariate independence, the uncalibrated MaxEnt reconstruction (red) aligns with the ground-truth distribution (gray), confirming that the conditional independence assumption suffices for uncorrelated covariates. **(b) Chain mediation:** When *X*_1_ sequentially influences *X*_2_, the uncalibrated MaxEnt approach (red) structurally attenuates the reconstructed HRs toward the null. Calibrated MD-JoPiGo (green), integrating empirical joint priors, corrects this drift. **(c) Collider selection:** Conditioning on *C* induces spurious correlation between *a priori* independent covariates. The uncalibrated MaxEnt reconstruction (red) distorts risk estimates, whereas calibrated MD-JoPiGo (green) recovers the ground-truth distribution. **(d) Robustness of the MaxEnt assumption**. Intrinsic multivariable log-hazard errors (Δ*β*) for synthetic cohorts (*N* = 400, 50 Monte Carlo iterations) reconstructed under the conditional independence prior (*P*_11_ = *P*_1_*P*_2_), evaluated across prescribed Pearson correlations (*r* ∈ [0.0, 0.9]). To isolate structural distortion from sampling noise, Δ*β* reflects the paired absolute deviation from ground-truth coefficients using Firth’s penalized regression. The dashed line delineates the Causal Robustness Envelope (Δ*β* = ln 1.5). The uncalibrated assumption incurs negligible error for weakly correlated covariates (*r*≤ 0.4) but introduces substantial systematic error for highly collinear covariates (*r* ≥0.6). **(e) Topological bounding via Fréchet gaps**. The maximum systematic error (Δ*β*_sys_) at Fréchet bounds is plotted as a function of the 1D Fréchet gap size (red funnel, left axis). Shrinking gap sizes bounds the multivariable error, ensuring convergence within the Causal Robustness Envelope. Concurrently, the reconstructed HR of a weakly correlated anchor covariate (*X*_3_; blue line, right axis) remains robust across all gap sizes. This demonstrates MD-JoPiGo’s “variance firewall”, preventing prognostic misallocation without requiring prior knowledge of true correlations. Box plots indicate median and interquartile range (IQR); violins display the density distribution of HR estimates.

Under parallel independence, the MaxEnt prior matches the true data-generating mechanism, recovering ground-truth multivariable hazard ratios (HRs). However, clinical covariates inherently violate this independence. Under mediation, the uncalibrated MaxEnt assumption enforces artificial conditional independence, structurally attenuating the reconstructed HRs toward the null. Integrating empirical joint priors (e.g., baseline cross-tabulations) overrides this default. Calibrated MD-JoPiGo thereby corrects the coefficient drift, restoring valid prognostic weights.

Collider stratification introduces trial-specific selection bias. When independent variables jointly determine trial enrollment, the cohort is effectively conditioned on a collider. This unblocks previously closed causal paths, inducing spurious dependencies (Berkson’s paradox) between *a priori* independent covariates^23^. By failing to account for this enrollment-driven selection mechanism, the uncalibrated synthesis distorts multivariable risk estimates. Conversely, calibrated MD-JoPiGo anchors the synthesis to the observed joint distribution. While calibration cannot reverse the original trial’s selection bias, it ensures fidelity to the enrolled population’s data-generating process, preventing secondary topological distortion during individual patient data (IPD) synthesis.

We evaluated the operational limits of the MaxEnt assumption through a sensitivity analysis across prescribed correlations (Fig. 2d). Paired absolute deviations from ground-truth coefficients isolate structural distortion from sampling noise. For weak-to-moderate correlations (*r* ≤ 0.4), the uncalibrated assumption incurs negligible error; reconstructions remain within the predefined Causal Robustness Envelope (Δ*β <* ln 1.5). Imposing the MaxEnt prior on highly correlated covariates (*r* ≥ 0.6) exceeds this threshold, introducing substantial systematic error. This error, however, requires relatively symmetric marginal incidence. Extended boundary analysis (Supplementary Fig. 1a–d) demonstrates that high marginal asymmetry truncates the achievable correlation via Fréchet bounds. This topological truncation bounds the operational error space, confining the synthesis within the robustness envelope even at the maximum identifiable correlation. These limits dictate the MD-JoPiGo synthesis strategy: the uncalibrated prior suffices for weakly correlated covariates, whereas structural calibration is required to resolve unidentifiable confounding in highly collinear, symmetric scenarios.

Within these highly collinear regimes, we quantified the impact of structural unidentifiability by evaluating the maximum systematic error (Δ*β*_sys_) at the Fréchet bounds as a function of the 1D Fréchet gap size (Fig. 2e). Shrinking this gap bounds the multivariable error, ensuring convergence within the Causal Robustness Envelope. Concurrently, the reconstructed hazard ratio of a weakly correlated anchor covariate (*X*_3_) remains robust across all gap sizes. By preserving the marginal association between the anchor covariate and the survival outcome, this structural isolation functions as a “variance firewall”, preventing prognostic misallocation without requiring prior knowledge of the true correlation matrix.

### Synthesis and structural calibration of multidimensional profiles in chain mediation topologies

Single-arm clinical trials frequently rely on synthetic control arms to evaluate relative treatment efficacy. Constructing these controls, however, requires multidimensional clinical profiles to adjust for baseline confounding. Because published reports typically present high-dimensional cohort data as independent one-dimensional (1D) Kaplan-Meier curves, the underlying joint distributions of patient characteristics remain unobservable. To determine whether MD-JoPiGo can recover these profiles from isolated marginal summaries, we applied the framework to a reference lung cancer dataset (*n* = 228)^25^, emulating a published single-arm cohort. This dataset is characterized by a chain mediation topology, wherein advanced age influences overall survival through the mediating variable of ECOG performance status.

The default maximum entropy (MaxEnt) method of the MD-JoPiGo framework synthesized multidimensional clinical profiles from 1D-IPD sets reconstructed from overall and stratified Kaplan-Meier curves. These unconstrained synthetic trajectories were statistically concordant with the reference ground truth at the one-dimensional level (Female; log-rank *P* = 0.95, Fig. 3a). To ensure representativeness, the displayed examples reflect subcohorts with median log-rank *P*-values within their respective tiers. However, this concordance exhibited a dimensional gradient, declining across two-dimensional interactions (Male and ECOG PS 0-1; *P* = 0.85) and reaching a statistical boundary in sparse three-dimensional combinations (Male, Age *<* 65, and ECOG PS 0-1; *P* = 0.65). This attenuation of statistical alignment is quantified by the uncalibrated log-rank results (Fig. 3b, empty grey circles). Survival comparisons for all remaining subcohorts are provided in Supplementary Fig. 2.

**Fig. 3.**
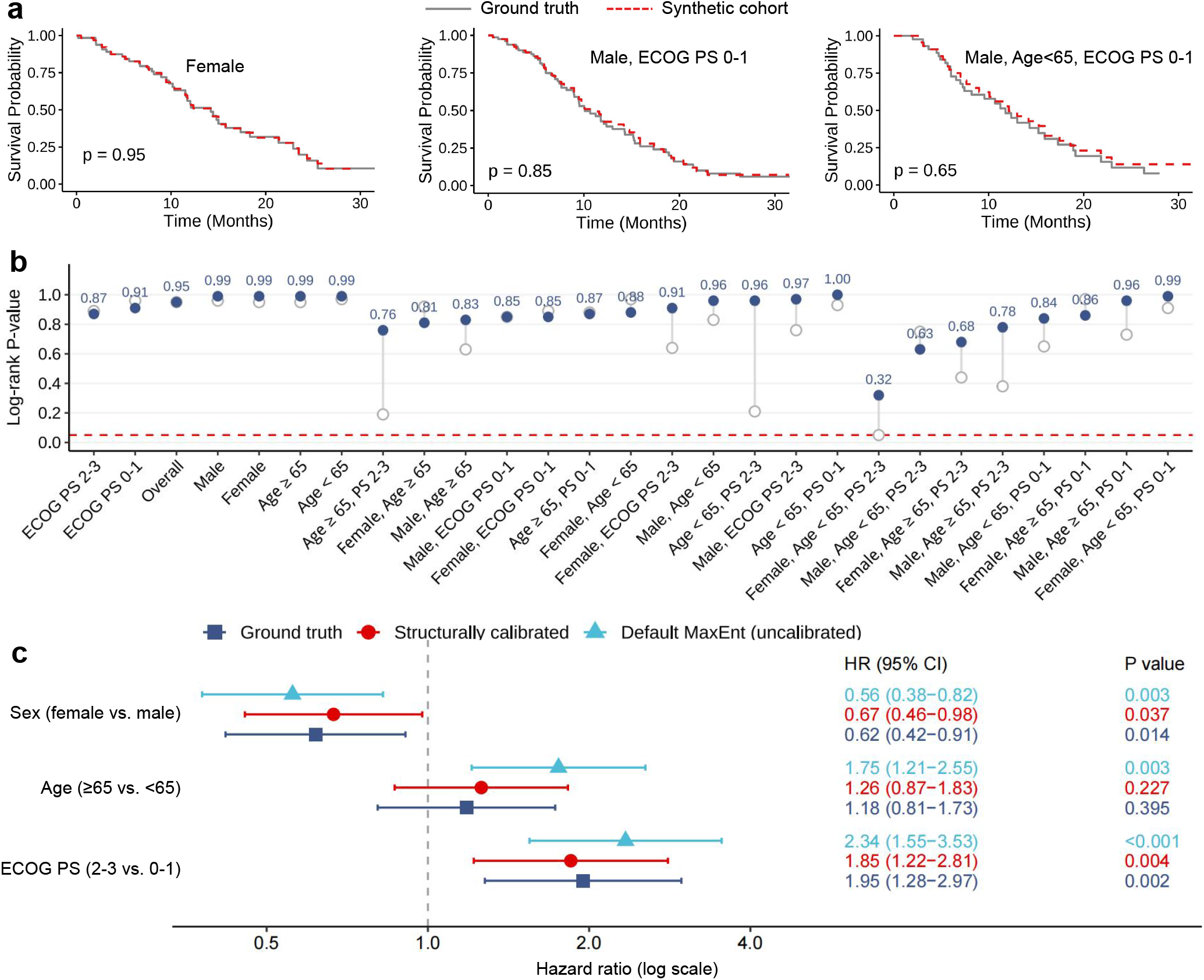
Structural calibration of synthesized multidimensional clinical profiles in an empirical lung cancer cohort. **a**, Kaplan–Meier survival trajectories of representative one-, two-, and three-dimensional subcohorts synthesized under unconstrained default maximum entropy (MaxEnt) assumptions. Displayed panels represent subcohorts with median log-rank *P*-values within each dimensional tier. **b**, Global assessment of subcohort survival concordance. Empty grey circles represent the uncalibrated synthesis, showing the attenuation of statistical alignment as combinatorial sparsity increases. Solid blue dots show the systematic recovery of survival concordance across unobserved interactions after incorporating a minimal structural prior (cross-tabulated proportions of age and ECOG performance status). The red dashed line marks the *P* = 0.05 significance threshold. **c**, Multivariable Cox regression analysis of independent prognostic weights. Uncalibrated synthesis (cyan triangles) exhibits localized coefficient drift, inflating the hazard ratio for age. Structural calibration (red circles) corrects this internal covariance deviation, restoring adjusted hazard ratios to the reference ground truth (dark blue squares). Error bars indicate 95% confidence intervals.

**Fig. 4.**
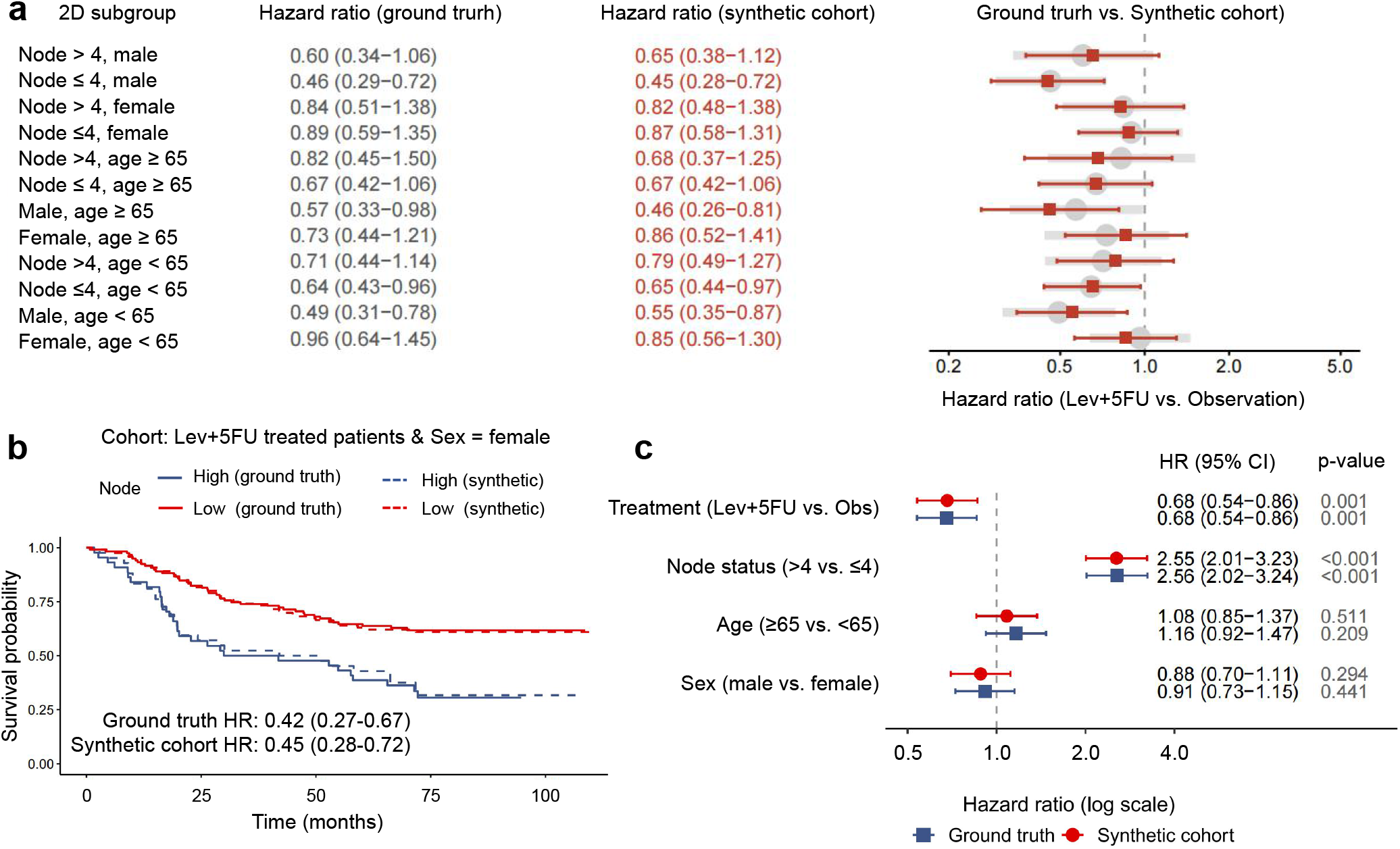
Intersectional treatment effect estimation in a synthesized clinical trial cohort. **a**, Forest plot evaluating the treatment effect (Hazard Ratio of Lev+5FU vs. Observation) across unobserved 2D interaction subgroups. The synthetic cohort (red squares) maintains statistical alignment with the reference ground truth (grey circles). **b**, Kaplan–Meier survival trajectories and corresponding prognostic HR for a granular 3D subcohort (Lev+5FU-treated female patients), comparing Node *>* 4 (blue) versus Node ≤4 (red). Solid lines denote the ground truth, while dashed lines represent the synthesized profiles. **c**, Multivariable Cox proportional hazards regression for the entire simulated trial cohort, verifying the independent prognostic weights of the assigned treatment and baseline covariates. Horizontal error bars indicate 95% CI.

To evaluate internal covariance within the synthesized profiles, we analyzed independent prognostic weights via multivariable Cox regression and risk prediction using clinical nomograms. While the adjusted hazard ratios (HR) for sex and ECOG performance status matched the reference data, unconstrained synthesis overestimated the prognostic effect of age (≥65 vs *<* 65). Because the default MaxEnt approach assumes conditional independence, it omits the inherent collinearity between advanced age and poorer performance status. This assumption redistributed shared prognostic variance, yielding an inflated HR for age (1.75 vs 1.18) and spurious statistical significance (*P* = 0.003 vs reference *P* = 0.395; Fig. 3c, cyan triangles). In the derived multivariable nomograms, age was correspondingly allocated disproportionate points, although the overall risk factor hierarchy (ECOG PS *>* Sex *>* Age) and survival prediction scales remained comparable to the reference models (Supplementary Fig. 3).

We recalibrated the synthesis using a minimal structural prior—the cross-tabulated proportions of age and ECOG performance status—to constrain the underlying joint distribution and correct the localized coefficient drift. This recalibration systematically improved survival concordance across unobserved two- and three-dimensional interactions, elevating the minimum subcohort log-rank alignment from *P* = 0.05 to *P* = 0.32 (Fig. 3b, solid blue dots; Supplementary Fig. 3). Within the multivariable Cox regression, the structural calibration resolved the internal covariance deviation: the adjusted HR for age (1.20; *P* = 0.351) reverted to the non-significant baseline, and the primary prognostic effects of sex and performance status remained consistent with the ground truth (Fig. 3c, red circles). To evaluate this topological sensitivity, we assessed all possible calibration combinations (Supplementary Table 1). The analysis demonstrated that synthesis fidelity relies on calibrating the specific causal dependency. Providing the functionally distinct Sex-ECOG prior did not correct the age-related bias (HR 1.64). In contrast, the minimal Age-ECOG prior eliminated the coefficient drift, achieving structural identifiability comparable to a fully calibrated topology. Minimal structural anchors thus counteract the variance redistribution inherent to unconstrained maximum entropy, generating structurally consistent joint distributions for precise trial emulation.

### Intersectional treatment effect estimation in a simulated clinical trial cohort

Identifying treatment effect heterogeneity across distinct patient subgroups is central to precision oncology^12^. While the joint distribution of baseline covariates determines specific treatment benefits, published randomized controlled trials (RCTs) typically collapse these dimensions into isolated marginal stratifications. This “dimensionality gap” often precludes the analysis of intersectional effects. Consequently, emulating trials through synthetic cohorts necessitates the reconstruction of high-dimensional profiles directly from these aggregated marginal summaries.

To assess the real-world robustness of MD-JoPiGo, we utilized a randomized controlled trial (RCT) dataset of stage III colon cancer (*N* = 929) comparing adjuvant levamisole plus 5-fluorouracil (Lev+5FU) versus observation^26^. To replicate the restricted data environments of published literature, we generated paired Kaplan–Meier plots from the reference IPD for the overall population and 1D-stratified subgroups defined by node status, age, and sex. These prognostic factors exhibit a topology of parallel independence (Scenario A), as node status, age, and sex operate as largely autonomous predictors of survival in this clinical context. These graphical summaries were digitized via KM-PoPiGo to reconstruct the individual patient data (IPD). Reflecting typical reporting limitations, the synthesis was constrained exclusively by sample sizes, at-risk tables, and censoring times, purposefully omitting specific event counts.

Treatment efficacy was then evaluated within unobserved 2D interaction subgroups, representing a clinically significant granularity for decision-making. Across these subcohorts, synthesized treatment hazard ratios (HR) remained consistent with the reference ground truth, with overlapping 95% confidence intervals (Fig. 5a). The framework recovered prognostic dynamics even in highly stratified subsets; for example, the survival trajectories and prognostic HR for node status among Lev+5FU-treated female patients aligned with actual data (Fig. 5b). This structural concordance was consistently observed across broader multidimensional stratifications defined by either demographic or clinical baseline characteristics (Supplementary Figs. 5 and 6). Multivariable Cox regression confirmed that the independent treatment effect and baseline covariate weights were preserved without significant coefficient drift (Fig. 5c), reflecting the lower collinearity among the trial variables compared to frailty-related indicators. MD-JoPiGo thus derives intersectional treatment effects and multi-variable trial dynamics directly from standard marginal inputs.

**Fig. 5.**
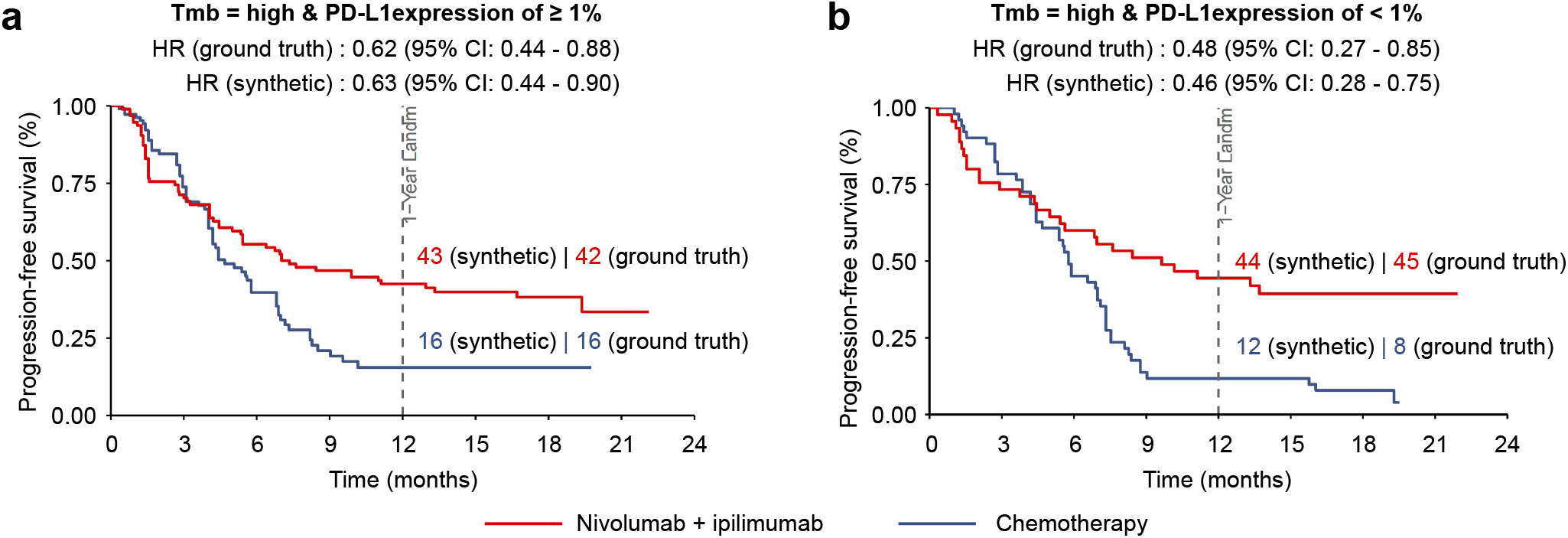
Validation of multi-arm cross-survival trajectories synthesized by MD-JoPiGo in multi-biomarker intersecting subgroups. **a**, Survival comparison between Nivolumab + Ipilimumab and Chemotherapy within the TMB-High and PD-L1 ≥ 1% subpopulation. The synthetic digital twin cohort demonstrated high concordance with the literature-derived ground truth. The ground truth hazard ratio (HR) is 0.62 (95% CI: 0.44–0.88), compared to the synthetic HR of 0.63 (95% CI: 0.44–0.90). The landmark 1-year survival rates for the synthetic cohort were 43% (Nivolumab + Ipilimumab) and 16% (Chemotherapy), strictly mirroring the reported clinical ground truth of 42% and 16%, respectively. **b**, Survival comparison within the implicitly reconstructed TMB-High and PD-L1 *<* 1% subpopulation. Despite the absence of explicit marginal survival constraints for the PD-L1 *<* 1% group, MD-JoPiGo accurately recovered the intersectional survival dynamics. The synthetic HR is 0.46 (95% CI: 0.28–0.75), aligning closely with the ground truth HR of 0.48 (95% CI: 0.27–0.85). The synthetic 1-year survival rates were 44% (Nivolumab + Ipilimumab) and 12% (Chemotherapy), consistent with the reported literature benchmark of 45% and 8%.

### Reconstructing latent intersectional efficacy from fragmented and asynchronous trial reports

Although previous sections validated MD-JoPiGo on complete single-study datasets, secondary analyses of published clinical trials present a distinctly more complex challenge. Critical biomarker stratifications are frequently reported across multiple publications over time, leading to asynchronous follow-up durations. Furthermore, biomarker data are often intrinsically fragmented. For example, while programmed death-ligand 1 (PD-L1) expression is typically assessed in the full intent-to-treat (ITT) population, tumor mutational burden (TMB) evaluation is strictly confined to a subset of patients with available tissue. Such spatial (population subsets) and temporal (follow-up) misalignments pose significant challenges to conventional summary-based reconstruction methods. To demonstrate how MD-JoPiGo harmonizes cross-publication data and resolves hidden intersectional topologies, we validated the framework using asynchronous reports from the CheckMate 227 trial.

We aggregated 1D marginal event-free survival (EFS) constraints from two independent publications. The overall EFS and evaluable TMB-stratified trajectories were reconstructed from published Kaplan-Meier (KM) images in an earlier report^27^. The full-population PD-L1 stratification curves (both ≥1% and *<* 1%) were obtained from a subsequent update with a longer follow-up period^28^. To resolve temporal discrepancies, we applied administrative right-censoring to truncate the extended PD-L1 trajectories at 24 months, ensuring aligned time horizons across all inputs. Although the earlier report [Insert Citation 1] provided the actual EFS outcomes for the joint biomarker subpopulations (e.g., TMB-High concurrent with PD-L1≥ 1% or *<* 1%), these intersectional trajectories were excluded from the reconstruction process. MD-JoPiGo synthesized the multidimensional clinical profiles using only the 1D margins, and the published joint outcomes served as a blinded ground-truth validation set.

Despite relying on an incomplete TMB subset and time-truncated PD-L1 populations, MD-JoPiGo reconstructed the multidimensional event-free survival (EFS) trajectories for both intersectional subgroups. Within the TMB-High and PD-L1 ≥1% subpopulation, the synthetic Nivolumab plus Ipilimumab versus Chemotherapy comparison yielded a hazard ratio (HR) of 0.63 (95% CI: 0.44–0.90), consistent with the literature-derived ground truth of 0.62 (95% CI: 0.44–0.88). The overall trajectories of the synthesized curves were visually consistent with the original published Kaplan-Meier plots^27^. Furthermore, the 1-year EFS rates for the synthetic cohort (43% vs. 16%) aligned with the clinical benchmarks (42% vs. 16%). For the TMB-High and PD-L1 *<* 1% subpopulation, the framework yielded a synthetic HR of 0.46 (95% CI: 0.28–0.75), comparable to the clinical benchmark of 0.48 (95% CI: 0.27–0.85), with synthesized curve trends also reflecting the published data. The reconstructed 1-year EFS rates in this subgroup (44% vs. 12%) approximated the reported literature values (45% vs. 8%).

To evaluate the framework under extreme data sparsity, we conducted a stress test by withholding the explicit PD-L1 *<* 1% marginal curves, providing the algorithm solely with the ITT and PD-L1 ≥ 1% margins (Supplementary Figure 7). In this scenario, MD-JoPiGo estimated the intersectional dynamics through residual topological space. The synthetic HR for the TMB-High and PD-L1 ≥ 1% cohort was 0.59 (95% CI: 0.41–0.85), with 1-year EFS rates of 46% and 20%. Within the unconstrained TMB-High and PD-L1 *<* 1% subgroup, the implicitly deduced HR was 0.48 (95% CI: 0.29–0.78), matching the clinical ground truth of 0.48. While the absolute 1-year EFS probabilities showed wider variances (36% vs. 4%) due to the relaxed constraint boundaries, the relative treatment hazard topology was preserved.

## Discussion

MD-JoPiGo reformulates the synthesis of multivariable clinical profiles from isolated 1D survival marginals as a deterministically bounded inverse problem^29^. Under weak correlation, the default maximum entropy (MaxEnt) assumption recovers ground-truth joint distributions with negligible error. However, highly collinear scenarios dictate structural calibration to restrict the unidentifiable parameter space and prevent prognostic misallocation. Estimating adjusted hazard ratios is fundamentally a high-dimensional feature attribution problem: it requires accurately distributing mortality risk across interacting covariates. By bounding the uncertainty of this marginal-to-joint synthesis, our framework ensures robust credit allocation^17^, enabling the reliable estimation of intersectional treatment effects routinely obscured in aggregate literature^30^.

Coefficient drift under high collinearity (*r* ≥ 0.6) stems from structural unidentifiability governed by the underlying causal topology^24^. In mediation or collider structures^31^, the uncalibrated MaxEnt prior mechanically severs inherent informational dependencies. By populating the synthetic cohort with unrealistic, “off-manifold” clinical profiles—such as implausible combinations of advanced age and pristine performance status—this unconstrained default misspecifies the data-generating mechanism^23^. This structural distortion triggers a critical breakdown in feature attribution: as covariate correlation increases, the Fisher information matrix approaches singularity, rendering the true joint distribution unidentifiable from marginal survival data alone^32^. The sharp error inflation at *r* ≥ 0.6 reflects this limit rather than stochastic sampling noise. Consequently, robust multivariable reconstruction mandates pre-evaluating covariate topology using Directed Acyclic Graphs (DAGs). This causal pre-screening dictates when targeted empirical priors must override the MaxEnt assumption to suppress spurious correlations and restore valid prognostic weights^23^.

Information theory dictates that reconstruction algorithms cannot synthesize unobserved joint variance. The identifiability of MD-JoPiGo is therefore governed by Fréchet inequalities. Simulations confirm that the maximum systematic error occurs when marginal probabilities are symmetric (e.g., *P*(*X*_1_) = 0.5, *P*(*X*_2_) = 0.5). Under these conditions, the Fréchet gap reaches its maximum, minimizing the constraints imposed by marginal trajectories. By computing this gap, the framework bounds the maximum systematic distortion (Δ*β*_sys_), deterministically identifying when supplementary clinical cross-tabulations are required to resolve the parameter space.

The framework’s “variance firewall” confines these topological distortions to the misspecified variables. Even at the Fréchet bounds of *X*_1_ and *X*_2_, the reconstructed hazard ratio of a weakly correlated anchor (*X*_3_) remains robust. This localization stems from two algorithmic safeguards. First, jointly permuting the anchor variable with survival outcomes (*T*, Δ) during simulated annealing preserves the true marginal association. Second, the penalized optimization landscape resolves the quasi-complete separation induced by mismatched “off-manifold” strata. By regularizing the objective function, it stabilizes the Fisher information matrix and blocks error contagion through the Hessian. Structural misspecification is thereby quarantined, ensuring that the prognostic credit allocation of the broader multivariable network remains intact.

Evaluating intersectional treatment efficacy drives precision oncology^33^, yet randomized controlled trial (RCT) publications routinely omit multivariable subgroup analyses^34^. The CheckMate 227^27,28^ reconstruction validates that when clinical biomarkers operate as parallel predictors, integrating asynchronous, fragmented 1D reports yields robust joint survival profiles. The computational generation of *in silico* data by coupling marginal distributions into joint probabilistic models has recently advanced multimodal single-cell omics^35^. Much like how summary-level statistics have been leveraged to unravel the genetic architectures of complex traits^36^, MD-JoPiGo translates this generative paradigm to the clinical domain. By synthesizing the intersectional clinical heterogeneity hidden within standard marginal summaries, the framework enables the construction of Synthetic Control Arms (SCAs)^37^ and external control cohorts. This capability transitions published clinical evidence from static aggregate reports to dynamic, individual-level trial emulation.

Validating synthetic trial emulation^38^ requires intersectional constraints. A single joint summary—such as the proportion of elderly patients with impaired performance status—proves sufficient to collapse the Fréchet gap and neutralize entropy-driven risk redistribution. Reporting joint counts for clinically collinear baseline characteristics is therefore not an administrative formality^39^, but a structural prerequisite for multivariable synthesis. Supplying these minimal empirical priors resolves structural unidentifiability while preserving individual-level privacy^40^.

Marginal information density defines the upper limit of synthesis fidelity. The CheckMate 227 reconstruction confirms that while relative treatment effects withstand data fragmentation, absolute survival probabilities depend on complete marginal constraints. Automating the extraction of clinical priors from trial corpora via Large Language Models (LLMs)^41^ provides a scalable path to generate these constraints. However, language-driven extraction requires deterministic bounding to prevent the propagation of spurious priors, anchoring the synthesis within verified topological boundaries. By constraining the structural unidentifiability between covariates, MD-JoPiGo recovers intersectional treatment dynamics previously restricted to proprietary datasets. This deterministic synthesis translates fragmented evidence into computable cohorts, providing the multivariable resolution required for individual patient data (IPD) meta-analysis and trial emulation.

## Methods

### Empirical datasets and clinical scenarios

To evaluate the structural identifiability and synthesis fidelity of the MD-JoPiGo framework across diverse causal topologies, we analyzed independent, publicly available oncology datasets. These datasets provide the complete multivariable joint distributions required to establish a ground truth reference for intersectional survival dynamics.

#### Colon cancer cohort (Parallel independence)

We analyzed data from a Phase III randomized controlled trial evaluating fluorouracil plus levamisole in stage III colon cancer (*N* = 929)^26^. This cohort provides a baseline for the *parallel independence* causal topology (Scenario A). In this dataset, baseline prognostic factors—such as sex, histological grade, and nodal involvement—act as independent predictors of disease-free survival. Because these variables lack severe structural collinearity, we can test the framework’s ability to resolve multivariable dynamics using default maximum entropy assumptions, without supplemental structural priors.

#### Lung cancer cohort (Chain mediation)

To evaluate the framework under interdependent structural constraints, we analyzed an advanced lung cancer dataset from the North Central Cancer Treatment Group (*n* = 228)^25^. This cohort represents a *chain mediation* topology (Scenario B). Clinically, advanced age contributes to physical frailty, such that age influences overall survival primarily through the mediating variable of ECOG performance status. This structural dependency typically induces localized coefficient drift during unconstrained synthesis. We used this dataset to demonstrate that incorporating a minimal structural prior (e.g., a single joint proportion of age and performance status) corrects this bias and recovers the reference multivariable profiles.

#### CheckMate 227 trial cohort (Fragmented and asynchronous data)

We analyzed asynchronous publications from the CheckMate 227 trial to evaluate the framework’s resolution of hidden intersectional topologies from fragmented real-world reporting. These asynchronous reports present spatial misalignments: programmed death-ligand 1 (PD-L1) expression was assessed in the full intent-to-treat population^28^, whereas tumor mutational burden (TMB) evaluation was confined to a patient subset^27^. Varying follow-up durations across the publications additionally introduced temporal discrepancies. To align the time horizons of the aggregated one-dimensional marginal event-free survival (EFS) constraints, we applied administrative right-censoring at 24 months. The published EFS outcomes for the joint biomarker subpopulations were explicitly excluded from the reconstruction process and reserved as a blinded ground-truth validation set.

For these empirical cohorts, the reference data were used to generate KM images for the overall population and 1D-stratified subgroups. These images were then digitized using the open-source KM-PoPiGo web tool (https://kmpopigo.github.io) to reconstruct one-dimensional individual patient data (1D-IPD) as standard CSV files. This process simulates the real-world workflow of extracting and synthesizing multidimensional clinical profiles from the dimensionally reduced survival summaries typical of published study.

### Extraction of one-dimensional individual patient data

We framed the extraction of 1D-IPD as a constraint-consistent inverse optimization problem. For this preprocessing step, we used the KM-PoPiGo tool to reconstruct individual time-to-event and censoring coordinates via joint global optimization. Although the tool can incorporate up to four empirical constraints—total sample size, number-at-risk tables, total event counts, and explicit censoring times—published stratified Kaplan-Meier plots rarely report exact event counts. To reflect this real-world data limitation, we excluded aggregate event counts from our reconstructions, relying entirely on sample sizes, at-risk tables, and censoring times. The resulting 1D-IPD sets, comprising (*t*_*i*_, *δ*_*i*_) tuples for each patient, provide the marginal boundary conditions for the multidimensional synthesis.

### Multidimensional joint distribution reconstruction

#### Problem Formulation

Consider the reconstructed IPD derived from the overall KM curve, comprising a cohort of *N* patients. Each patient *i* ∈ {1, …, *N*} is characterized by an observed tuple (*t*_*i*_, *δ*_*i*_), representing realizations of the time-to-event (*T*) and event indicator (Δ) random variables. The objective is to assign each patient *i* a multidimensional feature vector ***x***_*i*_ = [*x*_*i*,1_, *x*_*i*,2_, …, *x*_*i,D*_]^⊤^, which captures *D* clinical characteristics (e.g., disease stage, histology, and biomarker expression). Ultimately, we seek to reconstruct an *N*⨯*D* empirical patient-level state matrix **Θ** that preserves the underlying joint distribution *P*(***X***, *T*, Δ) and satisfies all marginal survival constraints reported in the source study.

#### Joint distribution estimation via maximum entropy

Prior to assigning individual clinical profiles, the joint probability distribution of the cohort must be estimated. Let 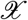 denote the state space spanning all potential combinations of the *D* clinical characteristics. For any specific state ***x*** ∈ 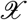, the objective is to estimate its joint probability *P*(***x***).

According to Jaynes’ principle of maximum entropy^42^, the most unbiased representation of the joint distribution is that which maximizes Shannon entropy, subject to known empirical constraints. This is formulated as a constrained optimization problem:

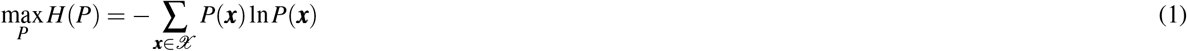

subject to the probability axioms:

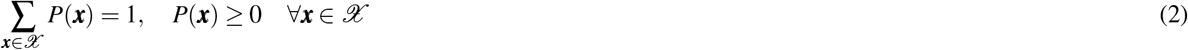

and the 1D marginal survival constraints reconstructed from published curves:

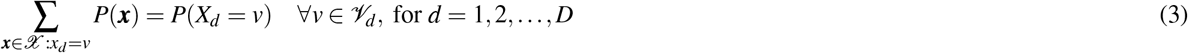

where *X*_*d*_ represents the *d*-th clinical variable, *𝒱*_*d*_ denotes the set of possible states for that variable (e.g., male, female), and *P*(*X*_*d*_ = *v*) is the observed marginal probability for each specific state.

The framework resolves the underlying causal topology by incorporating minimal structural priors. Under the baseline of *parallel independence*, optimization relies exclusively on 1D marginal survival constraints (Eq. 3), which inherently yields conditional independence under the maximum entropy objective. To overcome structural unidentifiability in *chain mediation* or *collider* topologies, the algorithm integrates supplemental constraints—such as a known joint probability *π*_*ab*_ between coupled variables *X*_*a*_ and *X*_*b*_—to anchor the reconstructed joint distribution:

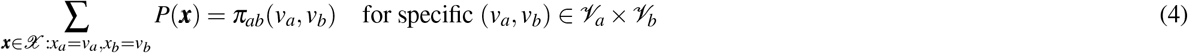

In practice, empirical marginals and structural priors extracted from published literature often contain minor numerical inconsistencies due to rounding or reporting artifacts. These slight deviations can render the strict intersection of hyperplanes defined by Eq. (3) and Eq. (4) mathematically empty, causing standard constrained solvers to fail. To ensure robust convergence and avoid algorithmic failure from empty feasible sets, we employ a quadratic penalty method. This approach relaxes the strict equality constraints into a soft penalty term, effectively expanding the feasible solution space while strongly penalizing deviations from the empirical priors.

The optimization objective is thus reformulated to maximize the regularized entropy:

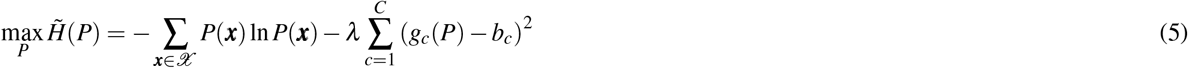

where *g*_*c*_(*P*) = *b*_*c*_ represents the *c*-th condition within the set of all *C* marginal and structural constraints, and *λ* is a hyperparameter imposing a large scaling penalty (e.g., *λ* = 10^4^) to enforce strict constraint satisfaction without inducing topological collapse.

Solving this unconstrained penalty optimization yields the optimal joint probability distribution *P*^*^(***x***). Scaling these probabilities by the total cohort size *N* determines the continuous target patient count. To establish the exact combinatorial boundary conditions for the individual-level label exchange, these continuous estimates are discretized into integer allocations *N*_***x***_ using the largest remainder method, guaranteeing that 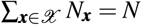 strictly.

#### Objective function formulation

The individual-level reconstruction is formulated as a combinatorial optimization problem. We define the objective function *J*(**Θ**) to quantify the discrepancy between the synthesized and reference marginal survival curves using the integrated squared error (ISE):

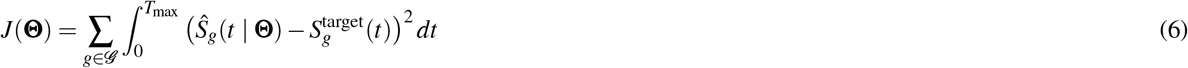

where:

- **Θ** is the *N* × *D* patient-level state matrix of clinical feature assignments.
- *𝒢* represents the set of all available 1D marginal subgroups (e.g., the overall cohort, male, female, age ≥ 65) serving as optimization targets.
- 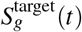 is the reference survival probability for subgroup *g* ∈ *𝒢*, derived from the initial 1D-IPD extraction.
- *Ŝ*_*g*_(*t* |**Θ**) is the Kaplan-Meier survival estimate computed from the synthesized cohort subset corresponding to subgroup *g* under the state matrix **Θ**.

In the computational implementation, to ensure high-speed evaluation during the simulated annealing iterations, this continuous integral is approximated over a uniformly discretized time grid across the interval [0, *T*_max_].

#### Optimization via simulated annealing

To minimize the non-convex objective function *J*(**Θ**), we employ simulated annealing (SA). The algorithm explores the search space by randomly selecting two patients and exchanging their multidimensional clinical feature vectors. This pairwise exchange mechanism ensures that both the predefined joint distribution constraints (*N*_***x***_) and the overall marginal frequencies remain strictly preserved throughout the optimization.

The acceptance probability for a candidate state transition **Θ** → **Θ**^′^ is governed by the Metropolis criterion:

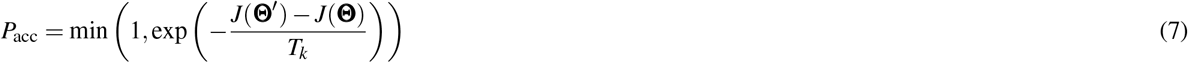

where *T*_*k*_ denotes the system temperature at iteration *k*, dynamically reduced according to a geometric cooling schedule *T*_*k*+1_ = *ηT*_*k*_. Based on empirical calibration, the algorithm is initialized at *T*_0_ = 2.0 with a cooling rate of *η* = 0.9998. The optimization executes for 50, 000 iterations, ultimately returning the best-found state matrix **Θ**^*^ that minimizes the observed objective error.

#### Downstream multivariable evaluation

Upon algorithmic convergence, the synthesized state matrix **Θ**^*^ serves as the foundation for downstream clinical evaluation. To assess the validity of the reconstructed intersectional profiles, the synthetic cohort is evaluated using multivariable Cox proportional hazards regression to estimate adjusted hazard ratios (aHRs):

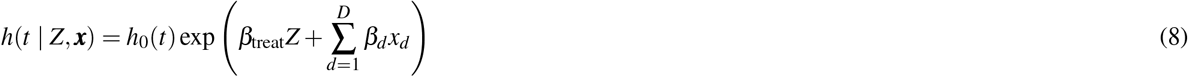

where *h*_0_(*t*) is the baseline hazard function, *Z* denotes the treatment indicator, and *x*_*d*_ represents the *d*-th component of the reconstructed baseline covariate vector ***x***, derived directly from **Θ**^*^. This step mathematically translates the computational output into standard clinical metrics for trial emulation and benchmarking.

### Downstream multivariable statistical inference

Upon algorithmic convergence, the optimized state matrix **Θ**^*^ yields the final synthetic multivariable cohort. To assess the prognostic validity of these reconstructed intersectional profiles, we perform downstream clinical evaluation using multivariable Cox proportional hazards regression. This standard survival analysis evaluates the adjusted hazard ratios (aHRs) for the synthesized individual patient data (IPD):

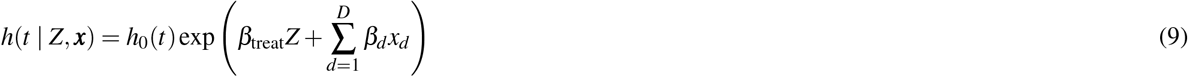

where *h*_0_(*t*) is the baseline hazard function, *Z* denotes the treatment indicator, and *x*_*d*_ represents the *d*-th component of the reconstructed baseline covariate vector ***x***, derived directly from **Θ**^*^. This inference step mathematically translates the multidimensional computational output into standard clinical metrics, establishing the foundation for cross-study benchmarking, synthetic control arm construction, and rigorous trial emulation.

### Simulation framework and causal topologies

To evaluate reconstruction fidelity, we conducted Monte Carlo simulations (*n* = 200 iterations per scenario), generating synthetic cohorts of *N* = 200 individuals each. For every simulated patient, two binary predictors (*X*_1_, *X*_2_) and a survival outcome (time-to-event *T* and censoring indicator Δ) were generated. Survival times were drawn from a Cox-Weibull model:

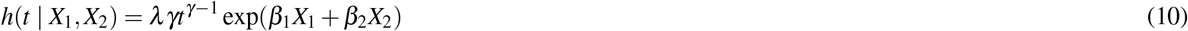

with baseline parameters set at scale *λ* = 0.01 and shape *γ* = 1.2. The target hazard ratios (HRs) were defined as HR_*X*1_ = 0.5 (*β*_1_ = ln 0.5 ≈ −0.693) and HR_*X*2_ = 0.75 (*β*_2_ = ln 0.75≈ −0.288). Independent right-censoring times were generated from an exponential distribution, calibrated to maintain an overall censoring rate of approximately 20%. We benchmarked the algorithm’s performance across three fundamental causal topologies:

- **Parallel independence (Scenario A):** *X*_1_ and *X*_2_ were modeled as independent and identically distributed (i.i.d.) Bernoulli trials (*p* = 0.5). In this baseline scenario, we evaluated the default MaxEnt mode (relying solely on 1D marginals) and compared its performance against a structurally calibrated reference (utilizing true joint frequencies). This comparison demonstrates that for independent predictors, the default MaxEnt assumption inherently matches the data-generating process, obviating the need for structural priors.
- **Chain mediation (Scenario B):** A directed dependency was introduced (*X*_1_ →*X*_2_), where *X*_1_ = 1 increased the probability of *X*_2_ = 1, inducing a Pearson’s correlation *φ* ≈ 0.5. For structural calibration, we provided MD-JoPiGo with the empirical 2 × 2 contingency table of *X*_1_ and *X*_2_. This prior information enables the algorithm to bypass the default independence assumption and allocate patient profiles according to the true joint probability.
- **Collider selection (Scenario C):** *X*_1_ and *X*_2_ remained independent in the source population, but inclusion in the cohort was conditioned on a collider *C* (*X*_1_ → *C* ←*X*_2_). Only individuals with *C* = 1 were retained in the analyzed sample, inducing an artificial negative correlation (Berkson’s paradox)^31^. Since the pre-selection baseline is inherently unobservable in empirical randomized controlled trials (RCTs)—which typically represent collider-conditioned populations—this simulation serves as the sole environment to evaluate synthesis fidelity under selection bias. Structural calibration was implemented by inputting the joint distribution observed within the selected sample, ensuring the reconstructed IPD preserves the trial’s internal multivariable dynamics and preventing secondary synthesis distortion.

For each topology, we compared the multivariable HRs derived from the ground truth sample against those estimated from the default MaxEnt and structurally calibrated reconstructions.

### Holographic Monte Carlo simulation framework

To rigorously evaluate the structural identifiability of MD-JoPiGo and preclude parameter-specific optimization artifacts, we developed a comprehensive “holographic” Monte Carlo simulation framework. Rather than testing isolated causal scenarios, this framework subjects the synthesis algorithm to an exhaustive spectrum of deterministic and latent topological parameters. We generated 200 independent synthetic cohorts (*N* = 400 patients each), simulating a three-node prognostic network (*X*_1_, *X*_2_, *X*_3_).

#### Deterministic marginals and unobserved causal topologies

To eliminate symmetric marginal bias and systematically evaluate how the size of the unobservable joint state space impacts reconstruction stability, the baseline marginal probabilities

*P*(*X*_1_) and *P*(*X*_2_) were deterministically modulated to traverse a uniform gradient of Fréchet bound widths (i.e., the mathematical gap between the theoretical maximum and minimum joint probabilities, ranging from 0.48 down to 0.02). Furthermore, to guarantee that the evaluation remains agnostic to underlying causal assumptions, a latent parameter *α* ∼ *U* (0, 1) was sampled for each cohort to randomly assign the true joint probability *P*(*X*_1_ ∩ *X*_2_) anywhere between mutual exclusivity (the Fréchet lower bound) and complete positive dependence (the Fréchet upper bound).

#### Weakly correlated anchor and variance firewall

A critical concern in multidimensional synthesis is the systemic propagation of estimation error: whether unidentifiable topological nodes improperly misallocate prognostic weights to other covariates. To evaluate the algorithm’s capacity as a “variance firewall,” we introduced a third prognostic anchor, *X*_3_. To simulate realistic clinical confounding, *X*_3_ was not rendered perfectly independent; rather, it was weakly entangled with *X*_1_ (e.g., *P*(*X*_3_ = 1 | *X*_1_ = 1) = 0.55, *P*(*X*_3_ = 1 | *X*_1_ = 0) = 0.35).

#### Stabilized disease dynamics and intrinsic error isolation

To strictly isolate topological distortion from baseline survival stochasticity (i.e., extreme sampling fluctuations or censoring artifacts), disease dynamics were governed by a stabilized multivariable Cox-Weibull kernel:

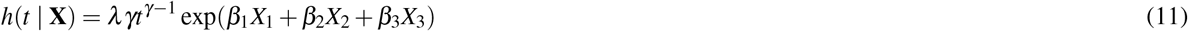

where the true log-hazard ratios were fixed at *β*_1_ = ln 0.5, *β*_2_ = ln 0.75, and *β*_3_ = ln(2*/*3). Both the Weibull scale (*λ* = 0.012) and shape (*γ* = 1.0) parameters, alongside a fixed administrative right-censoring rate, were locked to ensure uniform baseline information density. Consequently, paired relative errors derived from the simulated annealing reconstructions (100,000 iterations per cohort) reflect purely the mathematical distortion induced by structural misspecification.

### Evaluation metrics for structural identifiability

In traditional multivariable simulation studies, interaction *P*-values are frequently reported. However, relying solely on hypothesis testing in moderate-to-large samples (*N* = 400) is susceptible to the *P*-value fallacy, where statistically significant divergence may lack clinical relevance. Therefore, we shifted our primary evaluation metric to the absolute scale of clinical equivalence.

#### System maximum distortion and clinical equivalence

For each simulated cohort, we intentionally stripped the algorithm of any structural priors and forced it to reconstruct the multidimensional profiles under three extreme topological constraints: statistical independence, maximum positive correlation (the Fréchet upper bound), and maximum negative correlation (the Fréchet lower bound). The reconstructed cohorts were subsequently evaluated via Firth’s penalized multivariable Cox regression, which was crucial to mitigate sparse-data bias (e.g., quasi-complete separation) induced by extreme adversarial topologies.

To quantify synthesis fidelity and strictly isolate intrinsic algorithmic distortion from Monte Carlo sampling noise, we computed the maximum paired absolute deviation between the synthesized coefficients 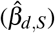 and the empirical ground-truth coefficients (*β* ^true^) of that specific matched cohort. Rather than selectively reporting isolated deviations, we tracked the *system maximum distortion* across the primary prognostic network:

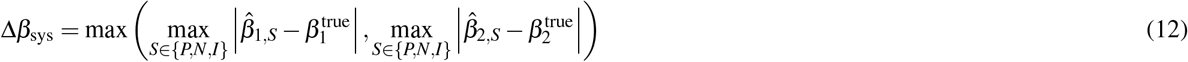

where *S* denotes the specific perturbation scenario (Positive, Negative, Independent). Structural identifiability was confirmed if Δ*β*_sys_ remained strictly bounded within a predefined Causal Robustness Envelope of ln(1.5) (representing a maximum tolerated hazard ratio shift of 50%), confirming that the synthesized multidimensional profiles were topologically locked by the 1D marginals irrespective of hidden dependencies.

### Full-spectrum sensitivity analysis of the independent prior

To systematically evaluate the robustness bounds of the maximum entropy (MaxEnt) assumption—which inherently assumes baseline predictors are conditionally independent in the absence of structural priors—we designed a full-spectrum correlation gradient test. We generated synthetic clinical cohorts sized representatively for oncology phase III trials (*N* = 400). To traverse the mathematical spectrum of the Pearson correlation coefficient (*r* ∈ [0.0, 0.9]), the marginal probabilities of both target binary predictors (*X*_1_ and *X*_2_) were fixed at 0.5. This symmetric 50*/*50 distribution was deliberately chosen as it mathematically maximizes the unobservable Fréchet gap size, thereby subjecting the algorithm to the absolute worst-case baseline uncertainty. The true underlying joint probability *P*_11_ was precisely modulated to achieve specific *r* values in increments of 0.1.

For each correlation level, we reconstructed the cohorts under the uncalibrated independence assumption (*P*_11_ = *P*_1_ × *P*_2_). To strictly isolate intrinsic algorithmic distortion from Monte Carlo sampling variance, the multivariable log-hazard error (Δ*β*) was computed as the paired absolute deviation from the empirical ground-truth coefficients of each matched cohort. Furthermore, we strictly employed Firth’s penalized likelihood Cox regression rather than standard maximum likelihood estimation (MLE). This penalization was crucial to mitigate sparse-data bias (e.g., quasi-complete separation or the Hauck-Donner effect). Such estimation pathologies occur when extreme adversarial reconstructions computationally force specific cross-tabulation microstrata (e.g., *X*_1_ = 0 ∩ *X*_2_ = 1) to near-zero event counts, which would otherwise induce infinite hazard ratios and artificially inflate the measured topological error. The maximum intrinsic multivariable error, 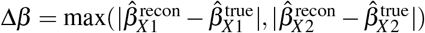, was benchmarked against a predefined Causal Robustness Envelope of Δ*β <* ln(1.5). This threshold represents the maximum acceptable prognostic deviation, ensuring that the induced error remains mathematically insufficient to overturn standard qualitative clinical inferences.

### Statistical analysis and evaluation metrics

All statistical analyses and IPD reconstructions were performed using the R statistical computing environment (version 4.3.3; R Foundation for Statistical Computing). Survival distributions were estimated using the Kaplan-Meier method, and multivariable prognostic dynamics were analyzed using the Cox proportional hazards regression model (via the `survival` package^43^). To quantify the performance of the MD-JoPiGo framework, we evaluated the synthetic cohorts against the original ground-truth data across two primary dimensions: marginal survival fidelity and multivariable intersectional consistency.

#### Statistical validation of survival concordance

The synthesis fidelity of the MD-JoPiGo framework was assessed via mathematical divergence metrics and statistical hypothesis testing. First, the deviation between the synthesized and reference marginal survival curves was quantified using the integrated squared error (ISE) for each subgroup *g* ∈ *𝒢*:

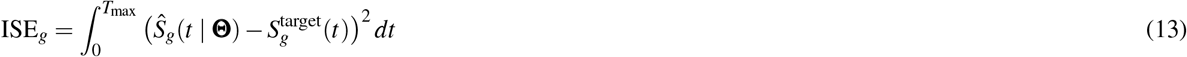

Second, to evaluate the statistical concordance between the synthesized multidimensional clinical profiles and the ground truth data, two-sided log-rank tests were employed across all stratified subpopulations. Rather than proving absolute equivalence, a resulting *P*-value *>* 0.05 was interpreted as an absence of statistically significant divergence, serving as a supplementary heuristic to the primary ISE metric to confirm synthesis fidelity.

#### Multivariable intersectional consistency

To assess the framework’s capacity to recover unobserved high-order interactions, the adjusted hazard ratios (aHRs) derived from the synthesized multidimensional clinical profiles were benchmarked against those obtained from the ground truth patient profiles. The relative error (RE) for each estimated log-hazard coefficient was quantified as:

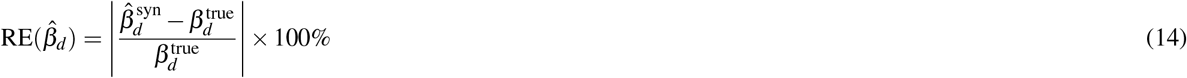

where 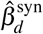 represents the coefficient estimated from the synthesized profiles, and 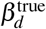 is the reference parameter. Furthermore, structural identifiability across varying causal topologies was evaluated by assessing whether the 95% confidence intervals (CIs) computed from the synthesized profiles successfully encompassed the ground truth multivariable aHRs.

## Data Availability

The MD-JoPiGo framework and its validation testing datasets are openly available at https://github.com/ZheqingZhu/mdjopigo. The empirical datasets used in this study (NCCTG lung cancer and Moertel colorectal cancer) are publicly accessible via the R 'survival' package (https://CRAN.R-project.org/package=survival).

## Code availability

The complete source code for the MD-JoPiGo framework, including the maximum entropy estimation and simulated annealing optimization modules, is open-source and publicly available on GitHub at https://github.com/ZheqingZhu/mdjopigo.

## Declarations

### Ethical approval

This study constitutes a secondary analysis of de-identified, publicly available data and summary statistics extracted from published clinical trials. No new human participants were recruited, and no clinical interventions were performed. Consequently, Institutional Review Board (IRB) approval and patient informed consent were deemed not applicable and were waived. All data synthesis and reconstruction procedures were conducted in accordance with the ethical principles outlined in the Declaration of Helsinki.

## Additional information

Supplementary Information is available for this paper.

