## Supplementary information for "Synthesizing multidimensional clinical profiles from published Kaplan–Meier images"

### Supplementary note

#### 1. Fréchet Truncation and Operational Boundaries under Asymmetric Marginal Distributions

While performance limits of the MaxEnt prior were previously defined under symmetric marginal incidence, clinical baseline characteristics—such as PD-L1 expression or specific genomic mutations—exhibit inherent asymmetry. To evaluate algorithmic robustness across these real-world scenarios, we extended the causal topology simulations to quantify the reconstruction error space under varying marginal asymmetry (major stratum incidence $P \in \{0.6, 0.7, 0.8, 0.9\}$).

The synthesis of joint distributions from 1D marginals is constrained by Fréchet inequalities[1]. For given marginal probabilities $P_1$ and $P_2$, the joint probability $P_{11}$ is bounded such that $\max(0, P_1 + P_2 - 1) \leq P_{11} \leq \min(P_1, P_2)$. This structural bounding truncates the maximum achievable Pearson correlation ($r_{\text{max}}$) between two binary covariates as marginal asymmetry increases.

We generated two-dimensional (2D) cohorts ($N = 400$) spanning prescribed correlations ($r = 0.0$ to $0.9$) and applied the uncalibrated MaxEnt assumption ($P_{11} = P_1 \times P_2$) to reconstruct multivariable profiles. Paired absolute deviations ($\Delta\beta$) of the reconstructed log-hazard ratios relative to the ground truth were computed to isolate structural distortion from sampling noise (Supplementary Fig 1).

Under moderate asymmetry (e.g., $P = 0.6$), the uncalibrated assumption mirrors the symmetric baseline: it yields negligible error for weakly correlated covariates ($r \leq 0.4$) but introduces substantial systematic distortion when correlations exceed the robustness threshold ($r \geq 0.6$). Under high marginal asymmetry (e.g., $P \in \{0.7, 0.8, 0.9\}$), the prescribed correlations are truncated at their respective Fréchet limits ($r_{\text{max}} \approx 0.43, 0.25$, and $0.11$).

This topological truncation intrinsically constrains the multivariable synthesis. Because the true underlying correlation cannot exceed these limits, the maximum reconstruction error is bounded. Under high marginal asymmetry, therefore, the uncalibrated synthesis remains confined within the predefined Causal Robustness Envelope ($\Delta\beta < \ln 1.5$). This analysis confirms that structural calibration is required when synthesizing highly correlated covariates with relatively symmetric distributions, whereas high asymmetry inherently restricts the magnitude of unidentifiable confounding.

#### 2. Survival matrix of overall and combinatorial subpopulations

This matrix evaluates the synthesized multivariable profiles across the overall population and all first- to third-order combinatorial strata defined by Sex, Age, and ECOG performance status. The synthetic cohort maintains statistical alignment with the ground truth, with boundary deviation ($P = 0.05$) restricted to the sparsest intersectional subset. This localized divergence defines the empirical limit of inferring complex joint distributions exclusively from marginal summaries.

#### 3. Evaluation of individual risk prediction using multivariable nomograms

To evaluate the impact of localized coefficient drift on clinical risk prediction, we constructed multivariable nomograms using the reference ground truth and the uncalibrated synthetic cohort. In the reference nomogram (Supplementary Fig. 3a), ECOG performance status accounts for the largest point allocation, followed by sex and age. The uncalibrated synthetic nomogram (Supplementary Fig. 3b) preserves this overall hierarchy (ECOG PS > Sex > Age) and maintains comparable mapping scales for 1-year and 2-year survival probabilities. However, consistent with the inflated hazard ratio observed in multivariable Cox regression, age ($\geq 65$) is allocated disproportionately higher risk points in the synthetic model. This inflation visualizes the clinical consequence of omitting the inherent collinearity between advanced age and poorer performance status under unconstrained default maximum entropy assumptions.

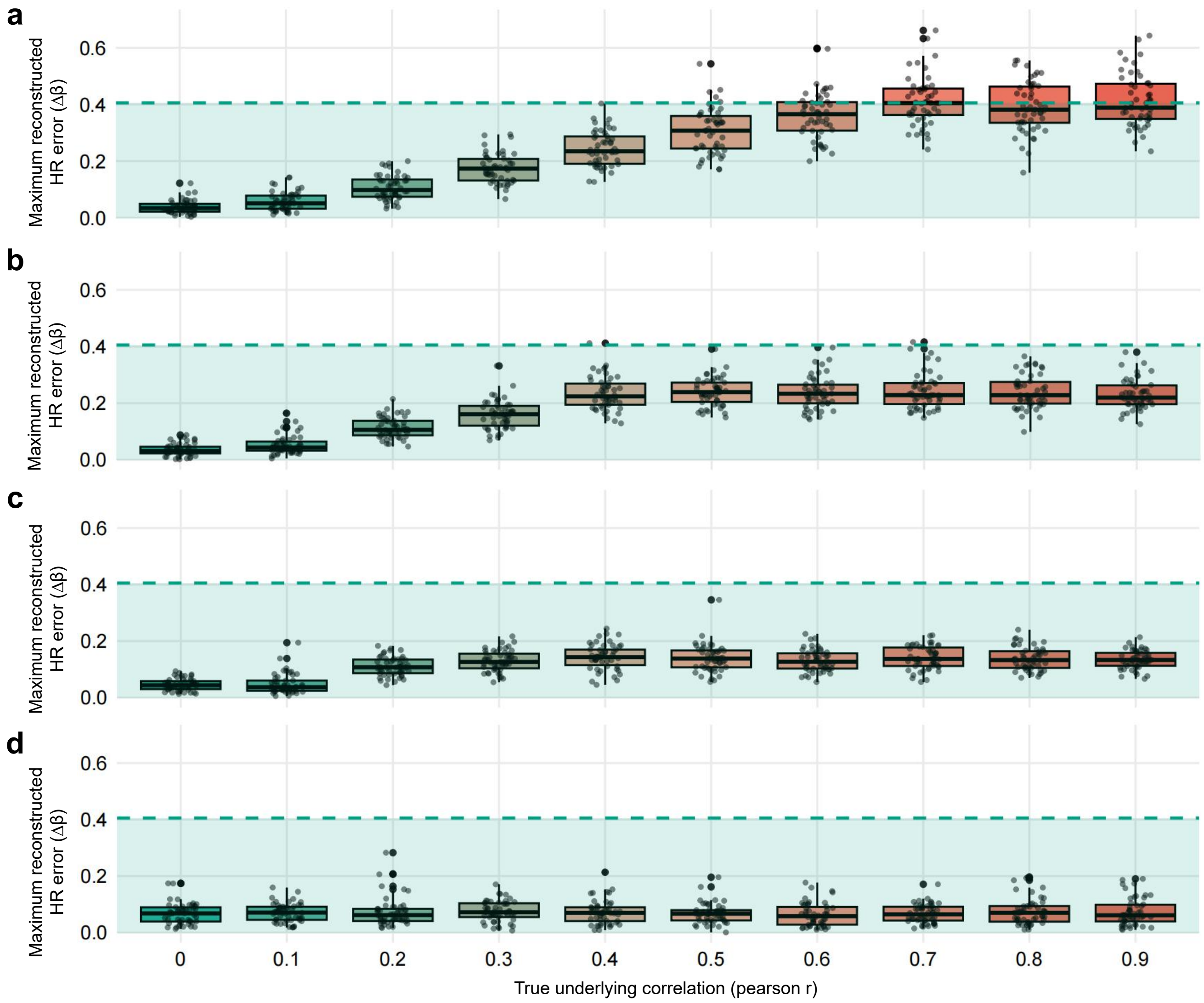

**Supplementary Fig. 1 Operational boundaries of the Maximum Entropy prior across asymmetric marginal distributions. a–d**, Maximum paired absolute deviations of reconstructed multivariable log-hazard ratios ($\Delta\beta$) across prescribed Pearson correlations. The uncalibrated MaxEnt assumption (conditional independence) was evaluated under four baseline marginal incidences (major stratum $P$): 0.6 (**a**), 0.7 (**b**), 0.8 (**c**), and 0.9 (**d**). Boxplots represent 50 Monte Carlo iterations per condition with a cohort size of $N = 400$. The dashed green line delineates the predefined Causal Robustness Envelope ($\Delta\beta = \ln 1.5$). Under moderate asymmetry (**a**), the uncalibrated synthesis incurs negligible error for weakly correlated covariates ($r \leq 0.4$) but introduces substantial systematic inflation at higher collinearity. However, as marginal asymmetry increases (**b–d**), the maximum correlation is truncated by Fréchet bounds ($r_{\text{max}} \approx 0.43, 0.25, 0.11$, respectively). This topological constraint inherently bounds the operational error space, preventing the reconstruction from exceeding the $\ln 1.5$ robustness threshold even under high identifiable confounding.

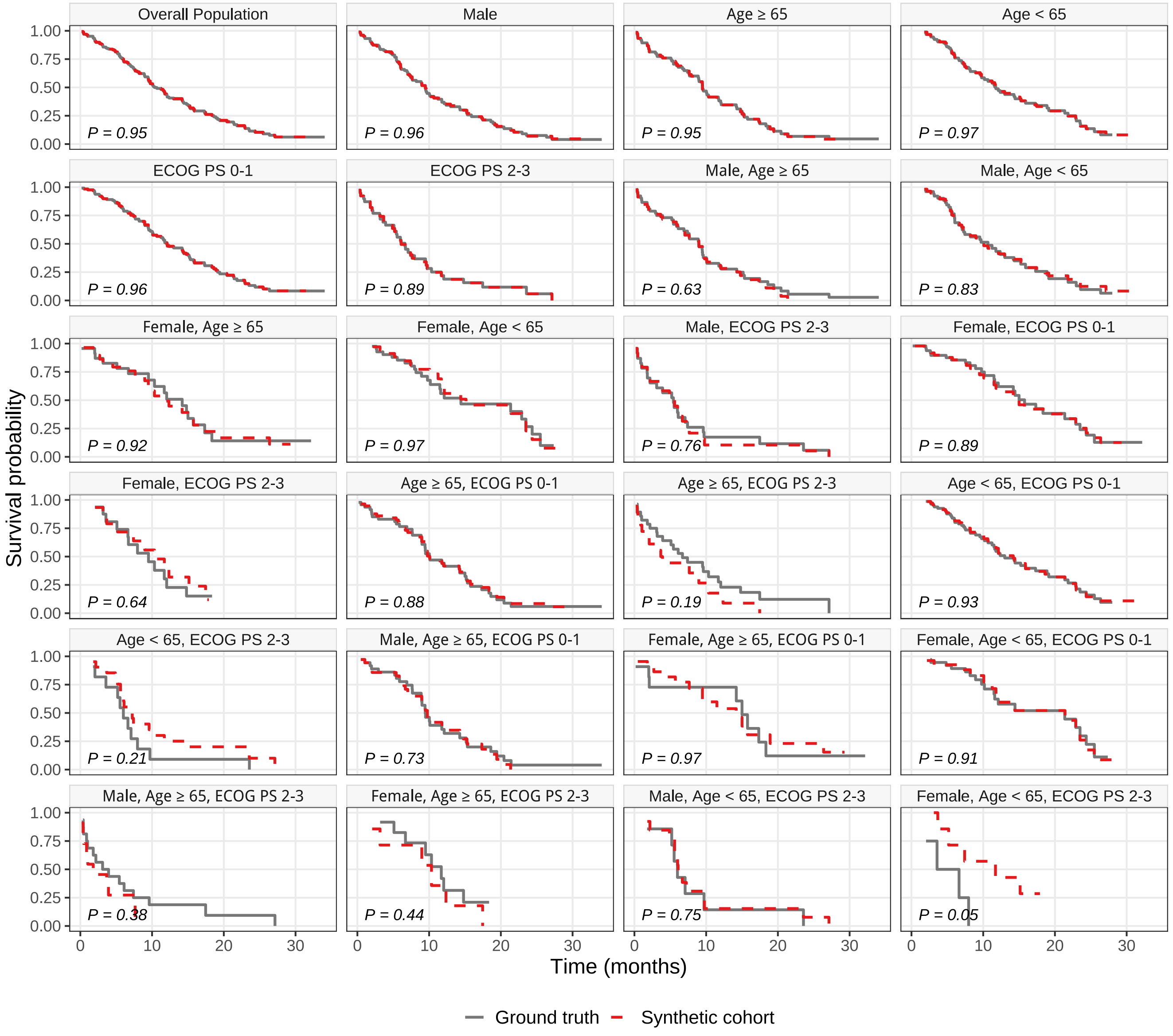

**Supplementary Fig. 2 | Systematic validation from marginal to joint survival distributions.** Kaplan–Meier survival trajectories comparing the reference ground truth (solid grey lines) with the uncalibrated synthetic cohort (dashed red lines). The matrix displays the overall population and all combinatorial subcohorts stratified by sex, age, and ECOG performance status. Log-rank $P$-values are provided for each panel to quantify the statistical alignment. While the synthesized profiles generally maintain concordance across lower-order strata, boundary deviations are observed in the most sparse three-dimensional subset (bottom right; $P = 0.05$).

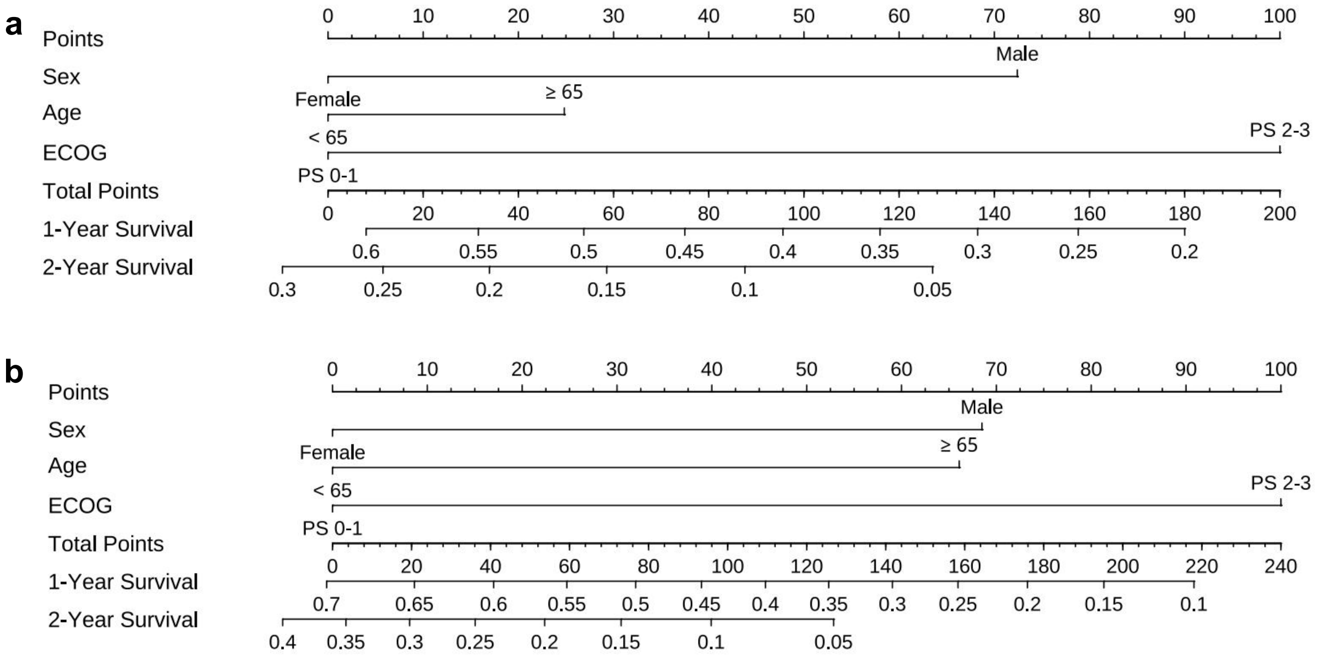

**Supplementary Fig. 3 Multivariable nomograms for individualized survival prediction. a**, Nomogram derived from the reference ground truth cohort. **b**, Nomogram derived from the uncalibrated synthetic cohort generated under default maximum entropy (MaxEnt) assumptions. Both models establish ECOG performance status as the dominant prognostic factor, followed by sex and age. However, the uncalibrated synthetic model allocates disproportionately higher risk points to the age covariate ($\geq 65$). This enlarged point allocation reflects the localized coefficient drift induced by the conditional independence assumption, which artificially redistributes shared prognostic variance from ECOG performance status to age.

#### 4. Survival matrix of combinatorial subpopulations following algorithmic recalibration

Supplementary to Fig. 4, this matrix presents the survival trajectories of the recalibrated synthetic cohort across all combinatorial stratifications of Sex, Age, and ECOG performance status. Integrating a single Age-ECOG joint proportion as an optimization constraint resolved the localized coefficient drift of the uncalibrated baseline. The recalibrated multidimensional profiles maintain statistical alignment with the reference ground truth across all demographic and clinical subcohorts. The boundary deviations previously noted in the sparsest intersectional strata are corrected, confirming that minimal structural priors yield structurally consistent joint distributions.

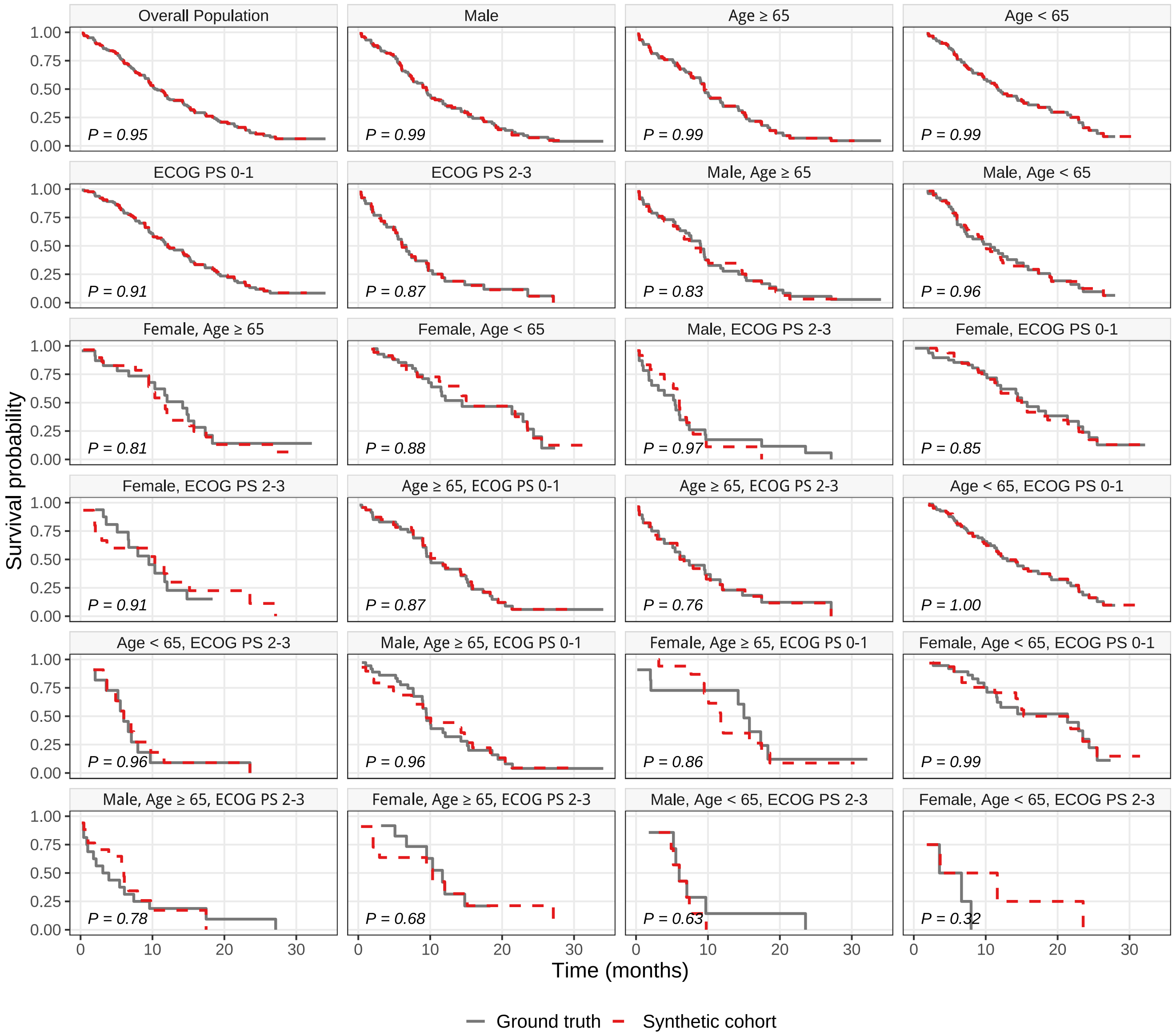

**Supplementary Fig. 4 Survival trajectories of the recalibrated synthetic cohort.** Kaplan–Meier curves comparing the reference ground truth (solid gray lines) and the recalibrated synthetic cohort (dashed red lines). The matrix presents the overall population alongside all single- and multi-variable subcohorts stratified by Sex, Age, and ECOG performance status. The synthesized profiles incorporate a minimal Age-ECOG intersectional constraint. Log-rank $P$-values in each panel show improved statistical alignment across all strata, eliminating the boundary deviations of the uncalibrated baseline (all $P \geq 0.32$).

### 5. Ablation analysis of calibration topologies

Supplementary Table 1 presents multivariable hazard ratios across all calibration topologies (Levels 0–3) for the lung cancer cohort. Under the default maximum entropy assumption (Level 0), unresolved collinearity between age and ECOG performance status artificially inflates the prognostic weight of age. Constraining the synthesis with the specific Age ↔ ECOG prior corrects this variance redistribution and recovers the ground-truth estimates. Calibrating functionally distinct pairs (e.g., Sex ↔ ECOG) does not resolve the bias. These results demonstrate that a minimal, targeted structural prior ensures structural identifiability without requiring the full joint distribution.

**Supplementary Table 1** Evolution of Multivariable Hazard Ratios for Age, Sex, and ECOG Across Comprehensive Calibration Topologies

| Topology Level | Calibration Constraints (Priors) | Age ($\geq$65 vs. <65) | Sex (Female vs. Male) | ECOG (2-3 vs. 0-1) |
|---|---|---|---|---|
| | | HR (95% CI) | HR (95% CI) | HR (95% CI) |
| **Ground Truth** | Original IPD Cohort | 1.18 (0.81–1.73) | 0.62 (0.42–0.91) | 1.95 (1.28–2.97) |
| **Level 0 (MaxEnt)** | None (Independent Assumption) | 1.75 (1.21–2.55) | 0.56 (0.38–0.82) | 2.34 (1.55–3.53) |
| **Level 1 (Single)** | Age ↔ ECOG | **1.26 (0.87–1.83)** | **0.67 (0.46–0.98)** | **1.85 (1.22–2.81)** |
| | Age ↔ Sex | 1.32 (0.91–1.92) | 0.65 (0.44–0.98) | 2.19 (1.45–3.31) |
| | Sex ↔ ECOG | 1.64 (1.14–2.36) | 0.52 (0.35–0.78) | 2.48 (1.64–2.36) |
| **Level 2 (Dual)** | Age ↔ ECOG + Age ↔ Sex | 1.14 (0.76–1.70) | 0.65 (0.44–0.95) | 1.81 (1.17–2.80) |
| | Age ↔ ECOG + Sex ↔ ECOG | **1.20 (0.82–1.77)** | **0.63 (0.42–0.92)** | **2.04 (1.31–3.16)** |
| | Age ↔ Sex + Sex ↔ ECOG | 1.31 (0.90–1.90) | 0.56 (0.38–0.82) | 2.19 (1.45–3.30) |
| **Level 3 (Full)** | All 3 Pairs (Complete Topology) | **1.26 (0.87–1.85)** | **0.61 (0.41–0.90)** | **2.11 (1.40–3.17)** |

Note: Results are derived from the multivariable Cox proportional hazards model.
HR: Hazard Ratio; CI: Confidence Interval.
↔ denotes a calibrated joint distribution prior between two specific clinical variables.

### 6. Multidimensional prognostic validation of synthetic cohorts

To evaluate whether the MD-JoPiGo framework preserves the prognostic relationships among intersecting patient characteristics, we conducted multidimensional stratified survival analyses within the Lev+5FU treatment arm. We compared subgroup-specific survival trajectories derived from the synthetic individual patient data against the ground-truth cohort. The cross-stratification analyses were defined by either baseline demographic subpopulations (age or sex; Fig. 5) or clinical severity strata (nodal involvement; Fig. 6). Across all evaluated intersecting strata, the survival distributions of the synthesized cohorts closely approximated those of the original data. The structural alignment of the Kaplan–Meier curves, alongside comparable hazard ratios (HRs) and overlapping 95% confidence intervals, demonstrates that the reconstruction process preserves the joint prognostic effects of multiple baseline variables without introducing structural distortions.

### 7. Evaluation of MD-JoPiGo under missing marginal information

In clinical literature, intersectional patient data are frequently obscured by incomplete reporting or asynchronous publication cycles. While MD-JoPiGo is designed to integrate all available 1D constraints, a critical requirement for its clinical utility is the capacity to maintain inferential integrity when certain marginal distributions are absent. To evaluate this 'information salvage' capability, we used the CheckMate 227 trial[2,3] to simulate an extreme data-sparsity scenario. A degraded input configuration was established by intentionally omitting the PD-L1 $< 1\%$ marginal EFS curves, which are typically required to anchor the reconstruction process. Under this setup, the algorithm was restricted to the ITT population and the PD-L1 $\geq 1\%$ subset. The objective was to determine whether the optimization engine could implicitly resolve the hazard topology of the unconstrained PD-L1 $< 1\%$ group (the 'hidden' margin) solely through the global constraints of the trial's overarching survival structure.

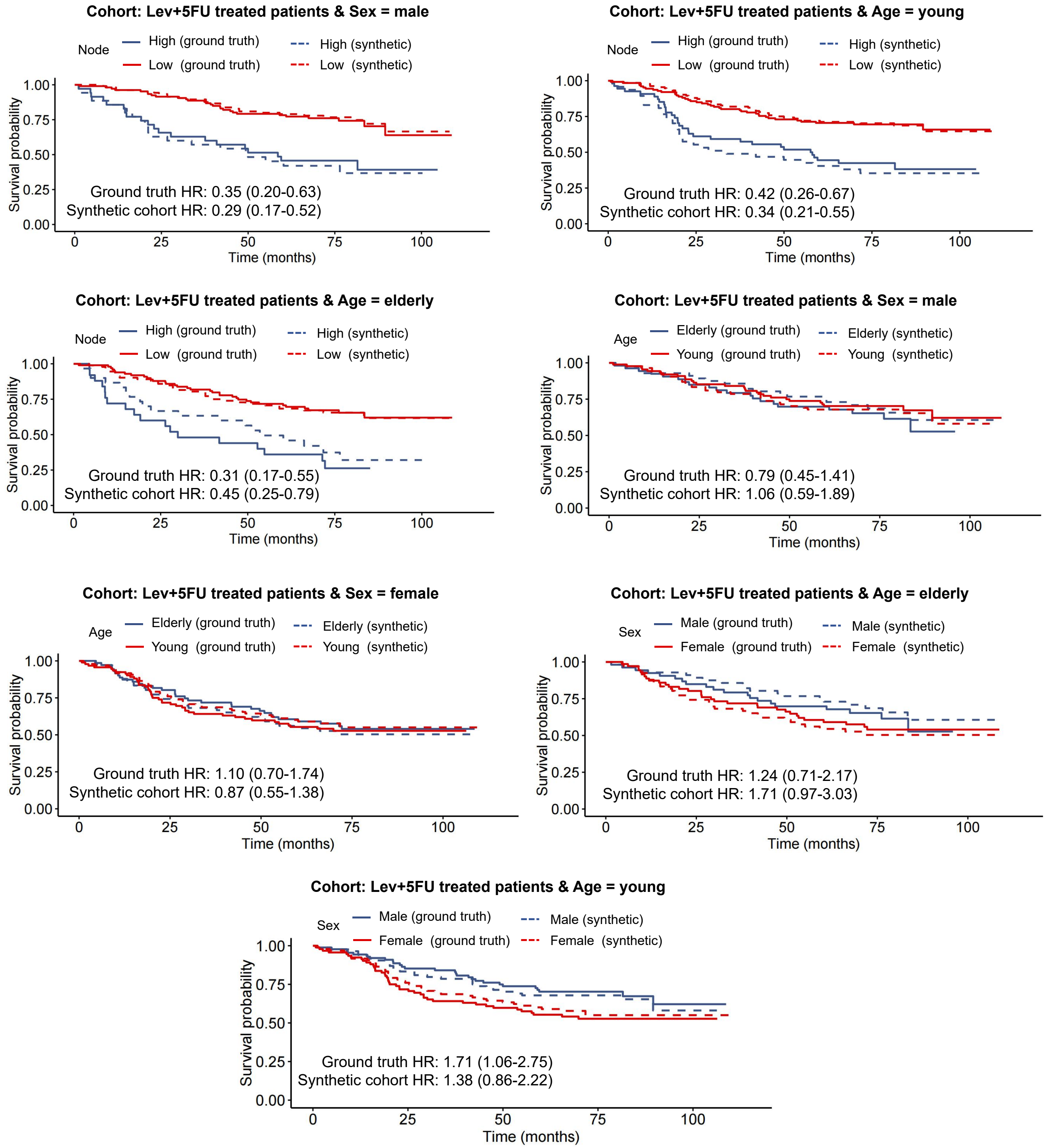

**Supplementary Fig. 5 Prognostic validation of synthetic cohorts within demographic subpopulations**
Subgroup-specific Kaplan–Meier survival trajectories comparing ground-truth individual patient data (solid lines) to MD-JoPiGo synthetic cohorts (dashed lines) in the Lev+5FU treatment arm. Patient cohorts were restricted to specific baseline demographic groups (age: elderly or young; sex: male or female) and further stratified by clinical features (for example, nodal involvement) or other demographic variables. The comparable subgroup-specific hazard ratios (HRs) and overlapping 95% confidence intervals demonstrate the framework's ability to preserve intra-subgroup survival distributions.

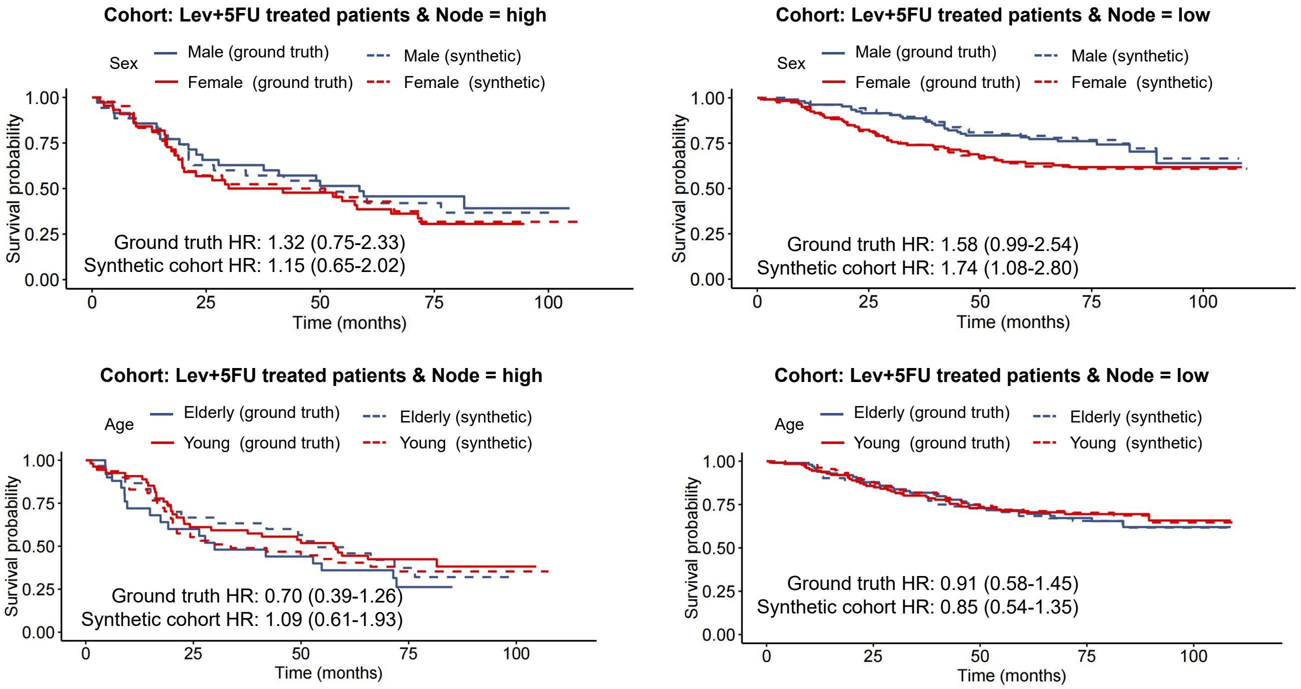

**Supplementary Fig. 6 Prognostic validation of synthetic cohorts within clinical severity strata.** Subgroup-specific Kaplan–Meier survival trajectories comparing ground-truth individual patient data (solid lines) to MD-JoPiGo synthetic cohorts (dashed lines) in the Lev+5FU treatment arm. Patient cohorts were restricted to specific baseline clinical statuses (nodal involvement: high or low) and further stratified by demographic variables (age and sex). The comparable subgroup-specific hazard ratios (HRs) and overlapping 95% confidence intervals demonstrate the framework's ability to preserve the prognostic impact of patient characteristics across different baseline clinical risk profiles.

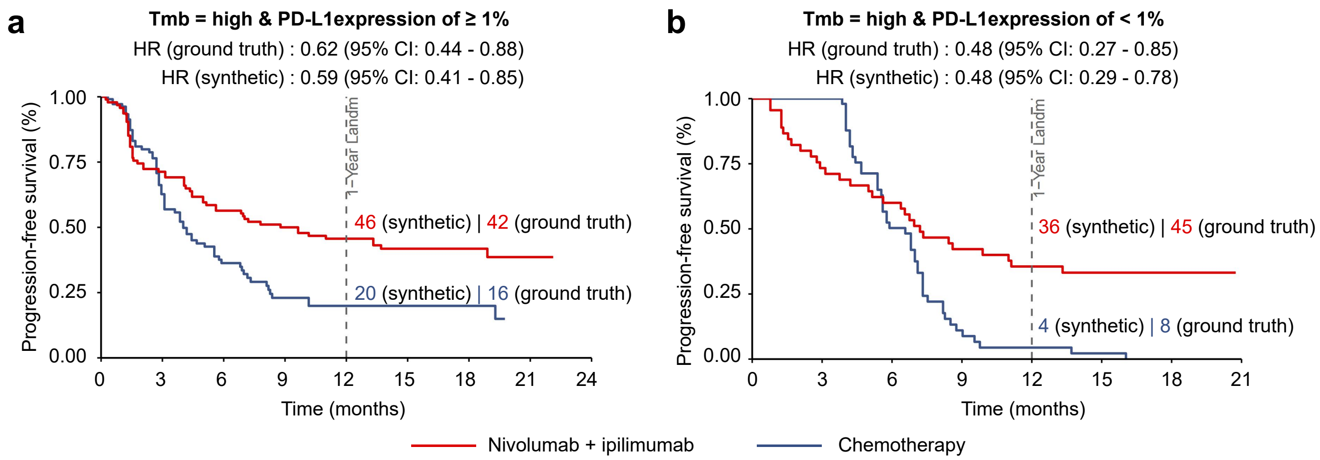

**Supplementary Fig. 7 Stress test of MD-JoPiGo under restricted marginal constraints using CheckMate 227 data. a**, Survival comparison between Nivolumab plus Ipilimumab and Chemotherapy within the TMB-High and PD-L1 $\geq 1\%$ subpopulation. In this analysis, the explicit PD-L1 $< 1\%$ marginal EFS curves were withheld from the framework. The synthetic hazard ratio (HR) was 0.59 (95% CI: 0.41–0.85), compared to the clinical benchmark of 0.62. The estimated 1-year EFS rates were 46% (95% CI: 37–57) for Nivolumab plus Ipilimumab and 20% (95% CI: 13–32) for Chemotherapy. **b**, Survival comparison within the unconstrained TMB-High and PD-L1 $< 1\%$ subpopulation. Despite the absence of direct 1D marginal constraints for this subgroup, the implicitly deduced HR was 0.48 (95% CI: 0.29–0.78), matching the clinical ground truth of 0.48. While absolute EFS probabilities at 1 year (36% [95% CI: 24–53] vs. 4% [95% CI: 1–17]) showed wider variances compared to the literature (45% vs. 8%) due to the relaxed optimization boundaries, the relative treatment hazard topology remained consistent with the reported trial outcomes.
